# The utility of homologous recombination deficiency biomarkers across cancer types

**DOI:** 10.1101/2021.02.18.21251882

**Authors:** Shiro Takamatsu, J.B. Brown, Ken Yamaguchi, Junzo Hamanishi, Koji Yamanoi, Hisamitsu Takaya, Tomoko Kaneyasu, Seiichi Mori, Masaki Mandai, Noriomi Matsumura

## Abstract

**Background:** Genomic alterations in *BRCA1/2* and genomic scar signatures are associated with homologous recombination DNA repair deficiency (HRD) and serve as therapeutic biomarkers for platinum and PARP inhibitors in breast and ovarian cancers. However, the clinical significance of these biomarkers in other homologous recombination repair-related genes or other cancer types is not fully understood.

**Results:** We analyzed the datasets of all solid cancers from The Cancer Genome Atlas and Cancer Cell Line Encyclopedia, and found that the association between biallelic alterations in the homologous recombination pathway genes and genomic scar signatures differed greatly depending on gender and the presence of somatic *TP53* mutation. Additionally, HRD cases identified by a combination of these indicators showed higher sensitivity to DNA-damaging drugs than non-HRD cases both in clinical samples and cell lines.

**Conclusion:** Our work provides novel proof of the utility of HRD analysis for all cancer types and will improve the precision and efficacy of chemotherapy selection in clinical oncology.

## Background

Homologous recombination repair (HRR) is one of the most accurate DNA repair mechanisms for DNA double-strand breaks (DSBs). Disruption of this mechanism (homologous recombination deficiency; HRD) leads to a high degree of genetic instability and accumulation of genetic mutations, thus playing an important role in the development and progression of cancer. Thus far, mutations in the *BRCA1* and *BRCA2* genes are considered principal drivers of HRD^1^; germline *BRCA1/2* mutation carriers more frequently develop *BRCA*-associated cancers, i.e., those of the ovaries, breasts, prostate, and pancreas^2^. Both germline and somatic *BRCA1/2* mutations are shown to be associated with high sensitivity to DNA-damaging drugs such as platinum, doxorubicin, and topoisomerase inhibitors^1^. After the discovery of synthetic lethality in *BRCA1/2* mutated cancers by PARP inhibitors in 2005^3,4^, several subsequent clinical trials validated this efficacy, leading to recent successive FDA approvals of PARP inhibitors for *BRCA*-associated cancers^5,6^.

In experimental studies, the suppression of HRR pathway signaling confers HRD properties in various cancers^7^, and thus gene mutations in the HRR pathway have been considered useful in predicting drug sensitivity associated with HRD. However, in a clinical setting, mutations other than *BRCA1/2* have not been sufficiently proven to be useful^8^. Moreover, the efficacy of PARP inhibitors in non-*BRCA*-associated cancers remains low, even in patients with *BRCA1/2* mutations^9-11^.

An alternative method for assessing HRD status is to detect characteristic patterns of genomic changes, referred to as genomic scar signatures. These indicators were developed in only the past decade, following remarkable improvements in sequencing techniques and the accumulation of large-scale multi-omics data^12,13^. As methods for scoring chromosome structural abnormalities due to HRD, the telomeric allelic imbalance (TAI) score^14^, the large-scale state transitions (LST) score^15^ and the loss of heterozygosity (LOH) score^16^ were developed, as well the sum of these scores which is titled the HRD score^17^. Furthermore, after the concept of mutational signatures was proposed^18^, a method for quantifying the characteristic mutational pattern in HRD tumors, generally referred to as mutational signature 3 (and referred to as Sig3 below), was developed^19^. In ovarian and breast cancers, platinum and PARP inhibitors have been found to be effective in tumors with high scores for these genomic scar signatures, even in the absence of *BRCA1/2* mutations^20^.

For carcinogenesis, tumor suppressor genes typically require not only a loss-of-function mutation in a single allele but also a biallelic alteration to fully induce a loss of the wild-type suppressive allele^21^. Recently, several methods for analyzing allele-specific copy number alterations from SNP genotyping array data or whole-exome sequencing data have been developed^22,23^, and combining these methods with genomic scar analysis, some studies have reported that the *BRCA1/2* mutation requires a loss of the non-mutated allele at the gene locus, termed locus-specific LOH, in order to possess its functional significance ^24,25^. However, these studies mainly focused on *BRCA1/2* mutations in *BRCA*-associated cancers; other cancers have been insufficiently analyzed. Furthermore, the association between zygosity status and genomic scar signatures in HRR pathway gene mutations other than *BRCA1/2* has yet to be investigated in detail. With the increasingly widespread use of gene panel and sequencing-based testing for personalized medicine, it is important to evaluate in full the significance of HRR pathway gene alterations and genomic scar signatures in a pan-cancer fashion.

Here, we comprehensively evaluate biallelic HRR pathway gene alterations and genomic scar signatures in all solid cancers from The Cancer Genome Atlas (TCGA) via an ensemble of analytical techniques. By including clinical information, we additionally examine the efficacy of DNA-damaging drugs in HRD cases. Not only is the clinical significance of HRD across cancer types clarified in a systematic way, but we also uncover a striking link between gender, TP53 mutations, and responses to DNA-damaging chemotherapeutic agents, which contributes to enhanced precision in personalized medicine based on HRD status.

## Results

### Correlations between HRR pathway gene alterations and genomic scar signatures

The HRD score^17^ was used as an indicator of chromosome structural abnormalities due to HRD. The HRD score’s underlying scores (TAI/LST/LOH) were strongly correlated (Additional file 1: Figure S1). Based on the literature^25^, we also calculated the Sig3 ratio (see Methods) as an additional indicator of somatic mutational patterns characteristic for HRD tumors. In what follows, the HRD score and Sig3 ratio are used as the measures of genomic scarring associated with HRD.

Using a combination of ASCAT^22^ and FACETS^23^ algorithms, allele-specific copy numbers at each locus of germline and somatic pathogenic variants in 29 selected HRR pathway genes were examined to determine whether they were accompanied by locus-specific LOH (see Methods for gene list). In short, the variants that were determined to have LOH by both algorithms, or to have LOH in one algorithm and to be unknown in the other, showed higher genomic scar scores than the other classification outcome groups (Additional file 1: Figure S2). Therefore, we defined these variants as those accompanying locus-specific LOH.

We examined the relationship between the genomic scar scores and the status of mutations with locus-specific LOH in HRR pathway genes; specifically, we considered the following six stratifications: germline(g)*BRCA1*, g*BRCA2*, gHRR (all HRR pathway genes excluding *BRCA1/2*, n=27, see Methods), somatic(s)*BRCA1*, s*BRCA2*, and sHRR (Fig. 1A,1B). When all of the cases were arranged in order of HRD score, mutations with LOH were significantly enriched in the high HRD score cases, while mutations without LOH were not; this trend was consistent in all six groups (Fig. 1A). The same trend was observed for the Sig3 ratio (Fig. 1B). Both the HRD score and Sig3 ratio were significantly higher in cases with LOH mutations than in cases with non-LOH mutations (Fig. 1A, 1B, box plot). For individual HRR pathway genes other than *BRCA1/2*, mutations with LOH in g*ATM*, g*BRIP1*, g*FANCM*, g*PALB2*, g*RAD51C*, s*ATM*, s*CDK12*, and s*FANCD2* were enriched in cases with high HRD score or Sig3 ratio, whereas mutations without LOH were not (Additional file 1: Figure S3). Mutations in these LOH-detected genes have been previously reported to be found in tumors with molecular features similar to *BRCA*-mutant tumors, known as “*BRCA*ness^1^.”

**Fig. 1.**
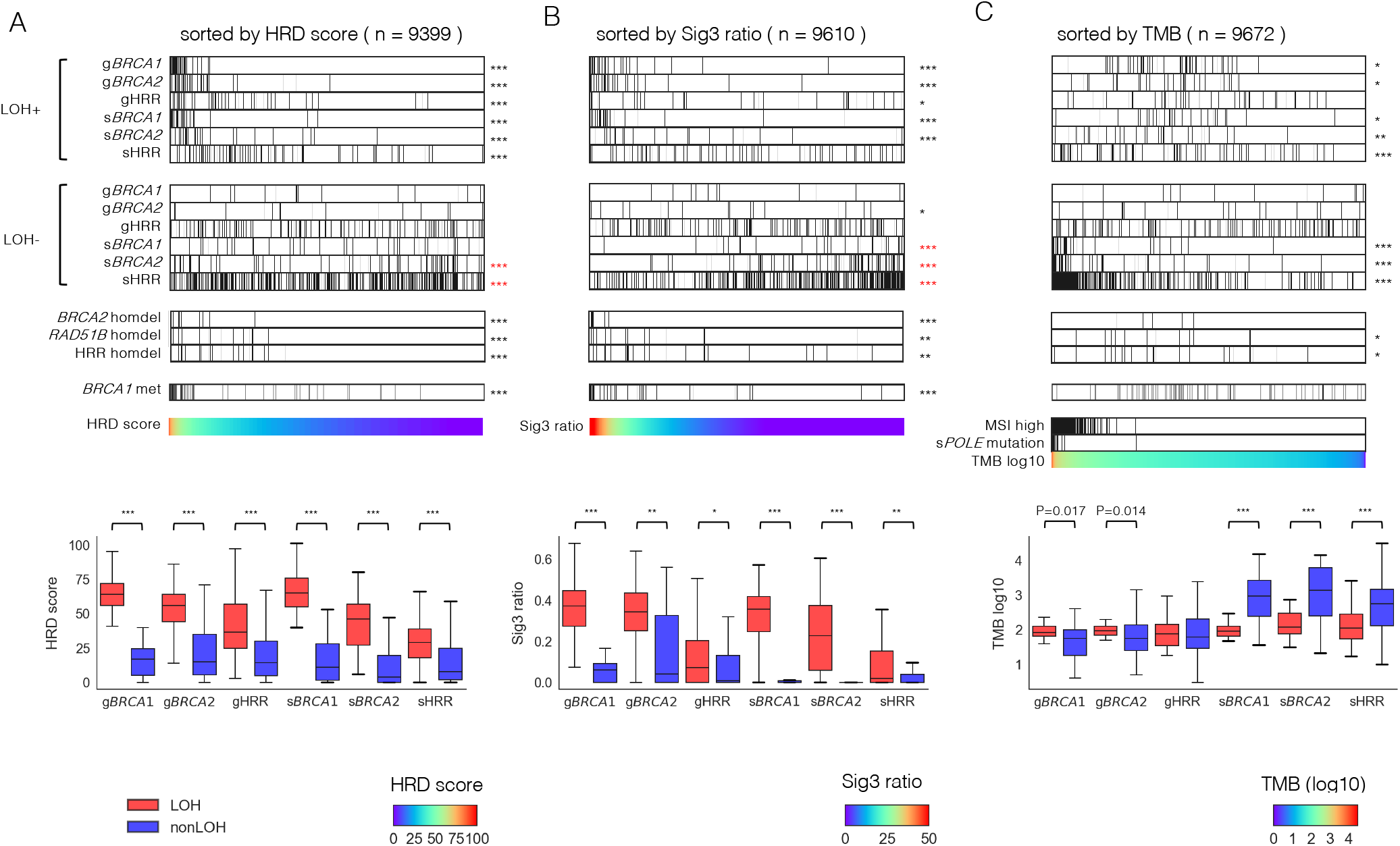
Association between HRR pathway gene alterations and genomic scar scores or tumor mutation burden (TMB). All samples were arranged in order of HRD score (A), Sig3 ratio (B), and TMB (C), respectively. The top and second panels show the distribution of mutations with and without locus-specific LOH, stratified by germline (prefix g) or somatic (prefix s), and *BRCA1, BRCA2*, or HRR pathway genes (except for *BRCA1/2*). The third panels show homozygous deletions in *BRCA2, RAD51B*, and HRR pathway genes (except for *BRCA1/2* but including *RAD51B*). The fourth panels show *BRCA1* methylation. The fifth panels contain the values used for ordering, and samples with MSI-high and somatic POLE mutation in C. The bottom box plots show comparisons of the scores between cases with and without locus-specific LOH in germline or somatic and *BRCA1, BRCA2* or other HRR pathway gene mutations, respectively. *, **, and *** stand for P < 0.01, P < 1× 10^−4^, and P < 1× 10^−6^ in the Mann–Whitney U test, respectively. Asterisks in red mean a negative correlation with the corresponding value.

Alternatively, when all of the cases were arranged in order of tumor mutational burden, mutations without LOH in s*BRCA1/2* and sHRR were strongly enriched in the hypermutated cases, including MSI-high and *POLE* mutated cases (Fig. 1C). Cases with somatic mutations without LOH had a remarkably high tumor mutational burden, suggesting that such mutations are neutral, known as “passenger,” mutations (Fig. 1C, box plot). In g*BRCA1/2*, cases with LOH mutations had relatively higher tumor mutational burden than cases with non-LOH mutations (g*BRCA1* p=0.017, g*BRCA2* p=0.014, Fig. 1C, box plot), which was consistent with previous reports that HRD tumors had moderately increased numbers of gene mutations^26,27^.

Homozygous deletions were annotated by combining two algorithms, ASCAT^22^ and ABSOLUTE^28^. Across all of the HRR pathway genes, cases determined to have homozygous deletion by both algorithms had higher genomic scar scores, whereas cases determined by only one algorithm had lower scores (Additional file 1: Figure S4A). Therefore, we defined homozygous deletion only when the results were matched in both algorithms. There were no homozygous deletions in *BRCA1*. Cases with *BRCA2* homozygous deletion had higher genomic scar scores and lower gene expression (Fig. 1A, B, Additional file 1: Figure S4C). Also, *RAD51B* homozygous deletion was found in 20 patients with high genomic scar scores and reduced gene expression (Fig. 1A,1B, Additional file 1: Figure S4B, 4C). Although *RAD51B* mutations have been reported in a few cases in breast cancer, their pathogenic significance remains controversial^29^. To the best of our knowledge, there have been no reports on homozygous deletion in *RAD51B*.

Regarding the association between the genomic scar scores with promoter methylation of HRR pathway genes, only *BRCA1* was significantly enriched in cases with a higher HRD score and Sig3 ratio (Fig. 1A,1B, Additional file 1: Figure S5). We, thus, used only *BRCA1* methylation in subsequent analyses.

### Differences in HRD status by cancer type

We next examined the mutation rate of HRR pathway genes and the frequency of locus-specific LOH for each cancer type. The TCGA ovarian cancer (OV) cohort contains only high-grade serous carcinoma^12^, which has the strongest HRD phenotype among the histopathological types of ovarian cancer. Consistent with previous reports^24,25,30^, *BRCA1/2* mutations tended to show higher LOH ratios in *BRCA*-associated cancers, i.e., ovarian, breast, pancreatic, and prostate cancers. However, the mean HRD score and Sig3 ratio were not particularly higher in breast, pancreatic, and prostate cancers compared to other cancers (Fig. 2A). Similarly, the frequency of biallelic alterations in HRR pathway genes (i.e., germline and somatic pathogenic mutations with LOH, or homozygous deletion) and *BRCA1* methylation by cancer type was not much higher in pancreatic and prostate cancers (Fig. 2B). After ovarian cancer, the next highest frequency of HRR alterations was observed in testicular germ cell tumors, in which most of the alterations were derived from *BRCA1* methylation. As previously reported, all of these cases were of the non-seminoma type^31^ and considered to have HRD with platinum/PARP inhibitor sensitivity^32,33^. The biallelic alterations in *ATM* were observed the second most frequently after *BRCA1/2* (Fig. 2B).

**Fig. 2.**
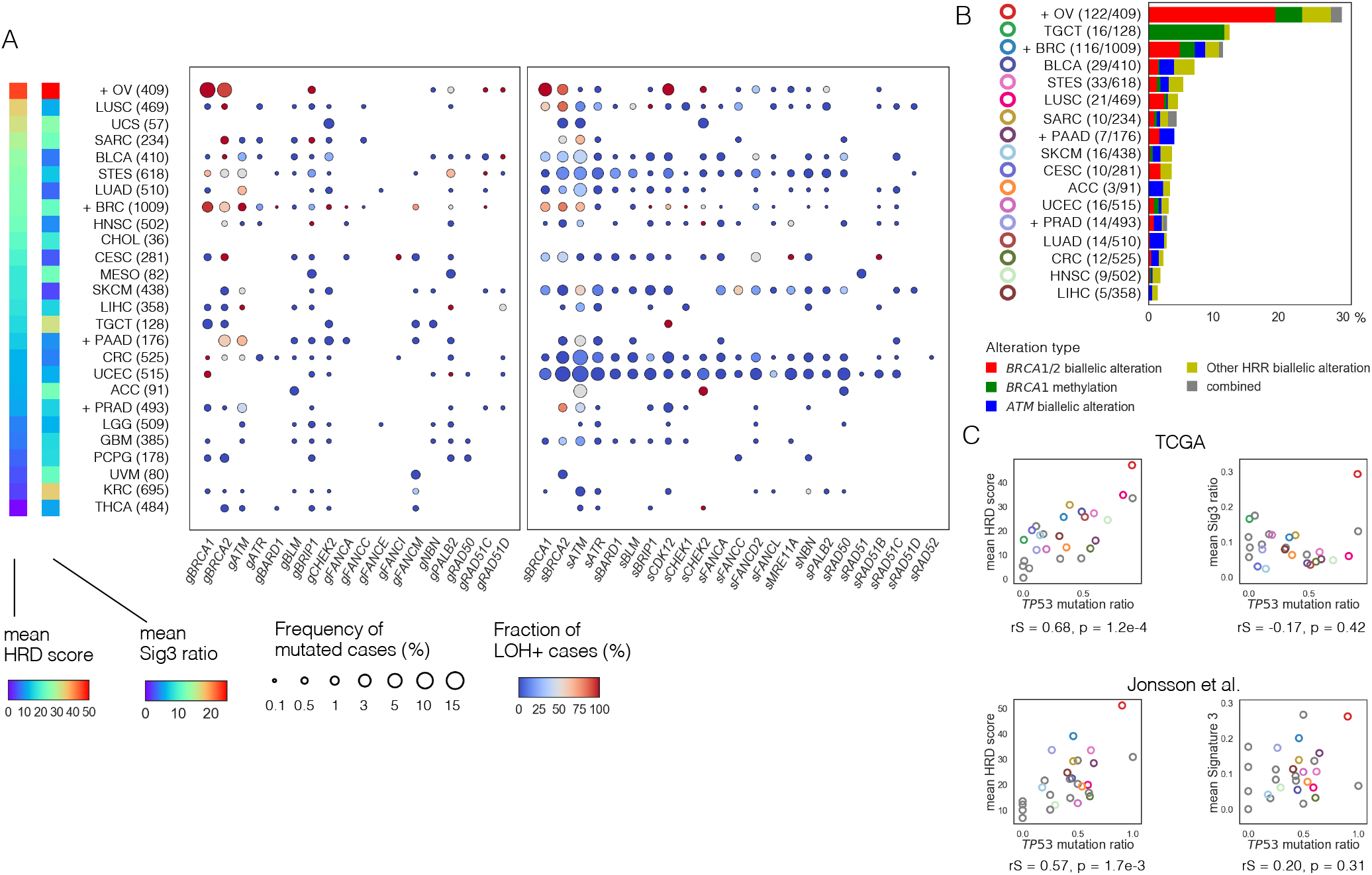
HRR pathway gene alterations per cancer type. A) Frequency of mutated cases and fraction of cases with locus-specific LOH for each HRR pathway gene per cancer type. Cancer types were arranged in order of the mean HRD score. The dot size indicates the frequency of mutated cases and the color indicates the fraction of involving locus-specific LOH in each gene. The prefix g in the gene name stands for germline and s for somatic. B) Types and proportions of biallelic HRR pathway gene alterations per cancer type. C) Correlation between *TP53* mutation ratio with mean HRD score (left) and mean Sig3 ratio (right) by cancer type. The upper panels are from the TCGA dataset and the lower from the Jonsson et al. dataset. The dot color matches the color of the cancer type shown in B, otherwise gray. rS and p represent the Spearman correlation coefficient and its p-value, respectively. A),B) Plus signs indicate belonging to *BRCA*-associated cancers. Numbers in parentheses represent the number of cases. Cancer names are abbreviated with reference to Broad GDAC’s cohort names (https://gdac.broadinstitute.org/); Adrenocortical carcinoma (ACC), Bladder urothelial carcinoma (BLCA), Breast invasive carcinoma (BRC), Cervical and endocervical cancers (CESC), Cholangiocarcinoma (CHOL), Colorectal adenocarcinoma (CRC), Glioblastoma multiforme (GBM), Head and neck squamous cell carcinoma (HNSC), “Kidney chromophobe, Kidney renal clear cell carcinoma, Kidney renal papillary cell carcinoma” (KIPAN), Brain lower grade glioma (LGG), Liver hepatocellular carcinoma (LIHC), Lung adenocarcinoma (LUAD), Lung squamous cell carcinoma (LUSC), Mesothelioma (MESO), Ovarian serous cystadenocarcinoma (OV), Pancreatic adenocarcinoma (PAAD), Pheochromocytoma and paraganglioma (PCPG), Prostate adenocarcinoma (PRAD), Sarcoma (SARC), Skin cutaneous melanoma (SKCM), Stomach and esophageal carcinoma (STES), Testicular germ cell tumors (TGCT), Thyroid carcinoma (THCA), Uterine corpus endometrial carcinoma (UCEC), Uterine carcinosarcoma (UCS), and Uveal melanoma (UVM).

Previous reports showed that *TP53* mutations were strongly associated with chromosomal instability across organs^34^ and elevated copy number change and HRD score^35^. Therefore, we hypothesized that differences in the genomic scar signatures between cancer types are related to the presence of *TP53* mutations. We resultingly found a strong positive correlation between the mean HRD score and *TP53* mutation ratio by cancer type (rS=0.68, p=1.2 × 10^−4^, Fig. 2C), whereas, there was no correlation between the mean Sig3 ratio and *TP53* mutation ratio. These results were similarly observed in the external data set based on various cancers reported by Jonsson et al.^25^

### Identification of pan-cancer HRD cases based on genomic scar signatures

The number of carriers of germline *BRCA1/2* variants is equal in women and men^36^ though the lifetime incidence of cancer is much higher in women^37^. Therefore, we hypothesized that the effect of HRD on cancer development would be gender-dependent. In addition, a previous report showed that the genome-wide LOH scores of tumors with biallelic *BRCA1/2* alterations differed greatly by cancer type and that prostate cancer, a male-specific tumor, was one of the lowest-scoring cancers^30^, although olaparib was shown to be effective in prostate cancer with *BRCA1/2* mutations^6^.

For the above reasons, we decided to evaluate the association between genomic scar scores and HRR pathway alterations separately by gender and *TP53* mutation status. The results showed that cases with *BRCA1/2* alteration, meaning germline and somatic *BRCA1/2* biallelic alteration plus *BRCA1* methylation, had the highest HRD score and Sig3 ratio in females with*TP53* mutations (Fig. 3A, left). The results were also similar in cases with HRR pathway gene alteration (HA cases), meaning germline and somatic biallelic alteration in HRR pathway genes and *BRCA1* methylation (Fig. 3A, middle). Also, in the Jonsson et al. dataset^25^, cases with biallelic *BRCA1/2* alteration had the highest HRD score and Sig3 ratio in females with *TP53* mutations (Fig. 3A, right). Furthermore, a similar trend was observed in cases with *BRCA1/2* alteration even when excluding *BRCA*-associated cancers (Additional file 1: Figure S6A). On the other hand, even in the group without HRR alterations, the HRD score was elevated with the presence of *TP53* mutation (Fig. 3A). These results indicate that stratification by gender and the presence of *TP53* mutation is needed to identify HRD cases based on the genomic scar scores.

**Fig. 3.**
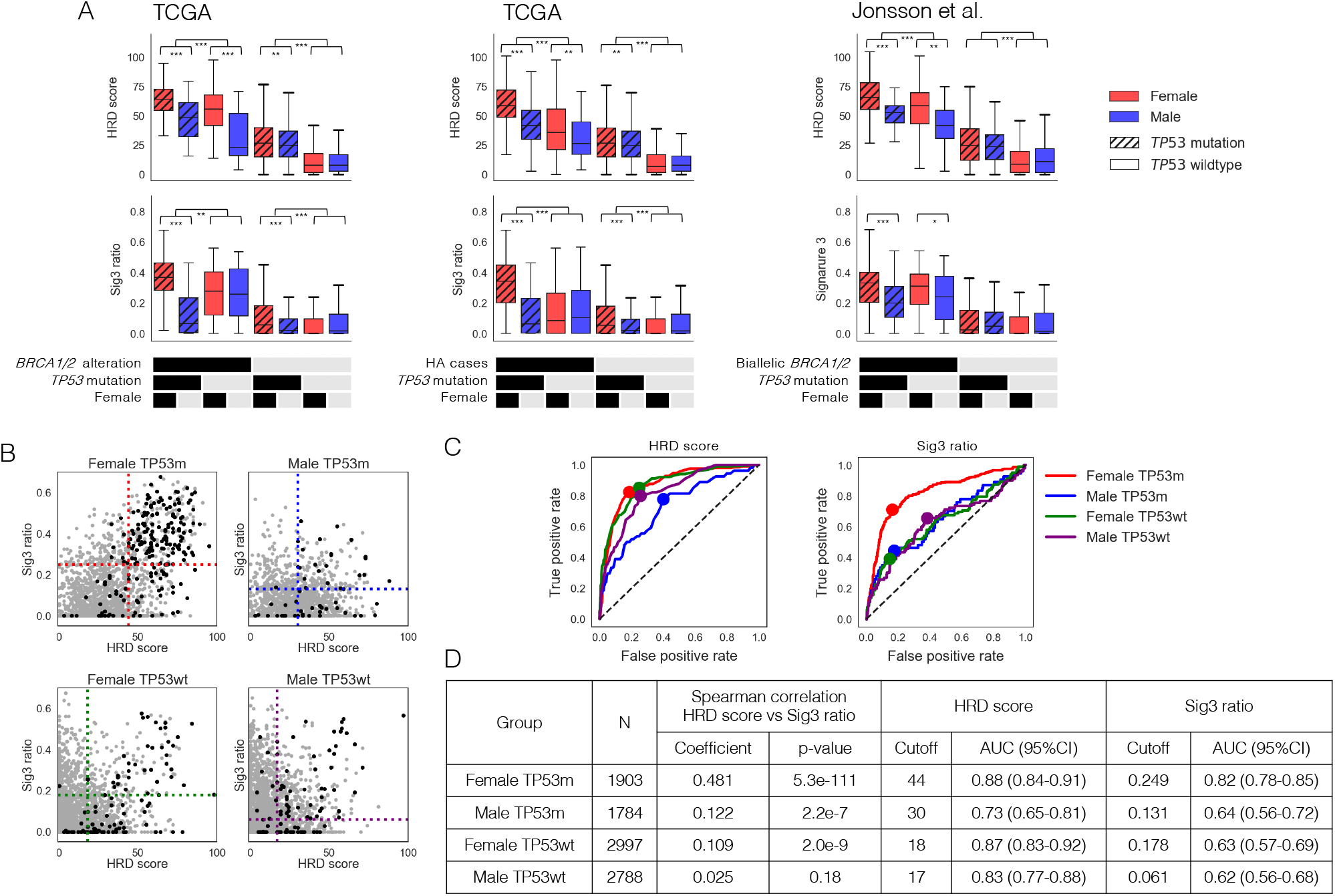
Identification of genomic scar high (GS) cases. A) Comparison of HRD score and Sig3 ratio in the eight groups divided by *TP53* mutation, gender, and (left) biallelic *BRCA1/2* alteration (including *BRCA1* methylation), (middle) biallelic HRR pathway gene alteration, (right) biallelic *BRCA1/2* alteration in Jonsson’s dataset. *, **, and *** stand for P < 0.05, P < 0.01, and P < 0.001 in the Mann–Whitney U test, respectively. B) Comparison of correlations between HRD score and Sig3 ratio and distributions of biallelic HRR pathway gene alterations among the four groups divided by gender and *TP53* mutation. Black dots represent the HRR pathway gene-altered cases and gray dots the other cases. The dotted lines represent the optimal cutoff values determined by the ROC curves in C. C) Differences in ROC curves and optimal cutoff values for HRR pathway gene alterations in HRD score and Sig3 ratio among the four groups. D) Spearman correlation coefficients between HRD score and Sig3 ratio, optimal cutoff values, and AUCs on the ROC curves among the four groups. TP53m and TP53wt indicate with and without *TP53* mutation, respectively.

The positive correlation between the HRD score and Sig3 ratio was strong in females with *TP53* mutations but weak in males with *TP53* mutations and females without *TP53* mutations and was non-significant in males without *TP53* mutations (Fig. 3B, rS=0.481, 0.122, and 0.109, 0.025, respectively). Similar results were still observed excluding *BRCA*-related cancers (Additional file 1: Figure S6B). After stratifying all cases into the four groups by gender and *TP53* mutation status, we examined the optimal cutoff values for determining HA cases by ROC curves in the HRD score and Sig3 ratio, respectively. We found that both the optimal cutoffs and the AUCs significantly differed among the four groups, with the highest values for both in females with *TP53* mutations (Fig. 3B, 3C, 3D). The cases that exceed both of the two cutoff values in each of the four groups were defined as genomic scar high (GS) cases.

### Gene expression analysis in GS and HA cases

The number of GS and/or HA cases was 341 in females with *TP53* mutations, 187 in males with *TP53* mutations, 233 in females without *TP53* mutations, and 357 in males without *TP53* mutations (Fig. 4A, Additional file 1: Figure S7). The rate of *BRCA1/2* alterations was highest in females with *TP53* mutations (Fig. 4A, 8.6%, 1.3%, 1.5%, and 1.2%, respectively, chi-square test p=6.5 × 10^−67^). The biallelic *ATM* alteration ratio was higher in the *TP53* wild-type group than in the *TP53* mutated group; *ATM* and *TP53* mutation were significantly mutually exclusive (Fig. 4A, 0.19%, 1.2%, respectively, chi-square test p=1.4 × 10^−7^).

**Fig. 4.**
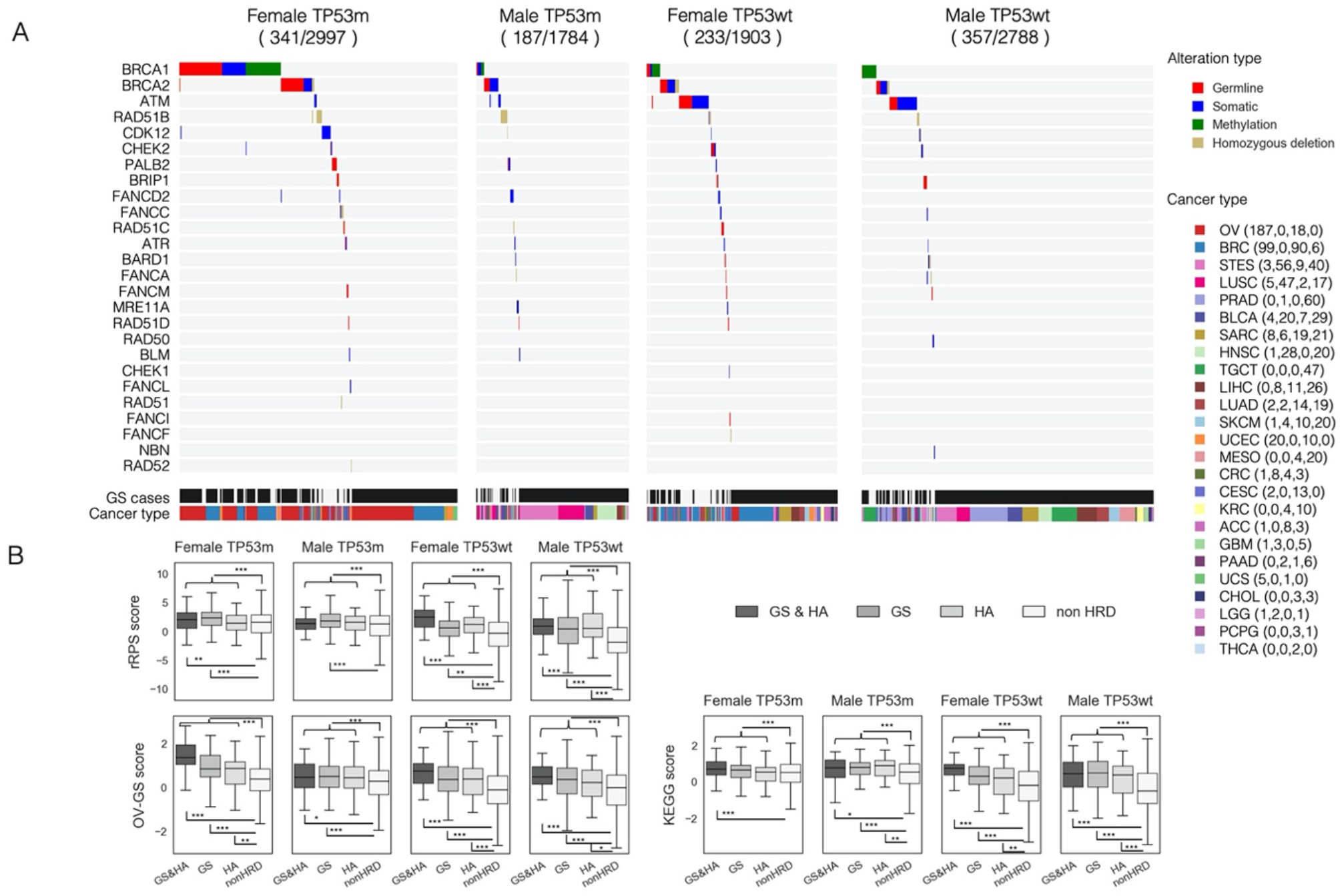
Characteristics of HRR pathway gene alteration (HA) cases and genomic scar high (GS) cases. A) Comparisons of the distribution of gene alterations and cancer types in HA and/or GS cases, after stratifying into the four groups by gender and *TP53* mutation. HRR gene alteration types and cancer types are color-coded as shown on the right side. Numbers in parenthesis next to cancer types represent the number of cases in the four groups, respectively. B) Comparisons of the gene expression-based HRD assessment scores between GS and/or HA and non-HRD cases. The p-values by the Mann–Whitney U test comparing GS and/or HA to non-HRD in each of the four groups were 2.1 × 10^−4^, 3.3 × 10^−4^, 1.0 × 10^−11^, and 3.4 × 10^−27^ in the rRPS score, 3.9 × 10^−23^, 4.2 × 10^−5^, 7.8 × 10^−15^, and 4.0 × 10^−15^ in the OV-GS score, and 7.3 × 10^−4^, 3.3 × 10^−7^, 2.6 × 10^−12^, and 3.7 × 10^−35^ in the KEGG score, respectively. *, **, and *** stand for P < 0.05, P < 0.01, and P < 0.001 in the Mann–Whitney U test, respectively. TP53m and TP53wt indicate with and without *TP53* mutation, respectively.

We next examined whether tumors defined as GS or HA cases had HRD properties in their gene expression profiles. By modifying the recombination proficiency score (RPS) in the previous report^38^, we calculated the reversed RPS (rRPS) score as a measure of HRD (see Methods). In addition, we identified differentially expressed genes (DEGs) from TCGA-OV cases based on both HRD score and Sig3 ratio (Additional file 1: Figure S8) and scored the enrichment of those DEGs in non-TCGA OV cases by the ssGSEA^39^ algorithm, referred to as the OV-GS score (see Methods). Furthermore, since the dysfunction of DNA repair mechanisms is reported to result in compensatory elevation of gene expression in that pathway^40^, using the KEGG homologous recombination pathway signature (hsa03440), we calculated the enrichment score of that signature by ssGSEA, referred to as the KEGG score (see Methods). All of the above three scores were significantly higher in GS and/or HA cases than in the other groups (Fig. 4B), and positively correlated with each other (Additional file 1: Figure S9). These results suggest that GS and/or HA cases, as defined by DNA alteration, have the characteristics of HRD in terms of gene expression profile. GS and/or HA cases are hereafter referred to as HRD cases.

In female cells, some of the molecules belonging to the DNA double-strand break repair pathway are reported to be involved in X-chromosome inactivation^41-43^. Therefore, we examined the association between HRD and X-chromosome inactivation. Expression of *XIST*, which is a non-coding RNA on the X chromosome with an essential function in X-chromosome inactivation, was significantly lower in female HRD tumors than non-HRD tumors (Additional file 1: Figure S10A). Additionally, the enrichment score of the whole X-chromosome gene set calculated by ssGSEA was significantly higher in female HRD tumors, whereas the score of a subset of genes reported to escape X-chromosome inactivation showed no significant elevation (Additional file 1: Figure S10A, see Methods). Also, we found that the global DNA methylation of the X chromosome was significantly lower in HRD tumors (Additional file 1: Figure S10A, see Methods). These differences were consistent even when *BRCA*-associated cancers were excluded (Additional file 1: Figure S10B). These results suggest that X-chromosome inactivation may underlie differences in HRD by gender.

### The association between chemotherapeutic agents and survival outcomes in HRD cases

Recently, it has been theorized that cancers with defects in a specific DNA damage repair mechanism are sensitive to drugs that target the exact mechanism^44^. According to this theory, HRD tumors can be considered susceptible to drugs that increase DNA damage or replication stress^45^. To investigate the drug sensitivity of HRD cases, all of the cases were divided into two groups: those with (n=2979) and without (n=6460) a treatment history of DNA-damaging agents, including alkylating agents, antibiotics, antimetabolites, platinum, and topoisomerase. While HRD cases had a significantly better overall survival than non-HRD cases in the group with DNA-damaging agent use, HRD cases in the group without treatment had a significantly worse outcome than non-HRD cases (Fig. 5A, log-rank test p=5.1× 10^−4^, 1.1 × 10^−10^, respectively).

**Fig. 5.**
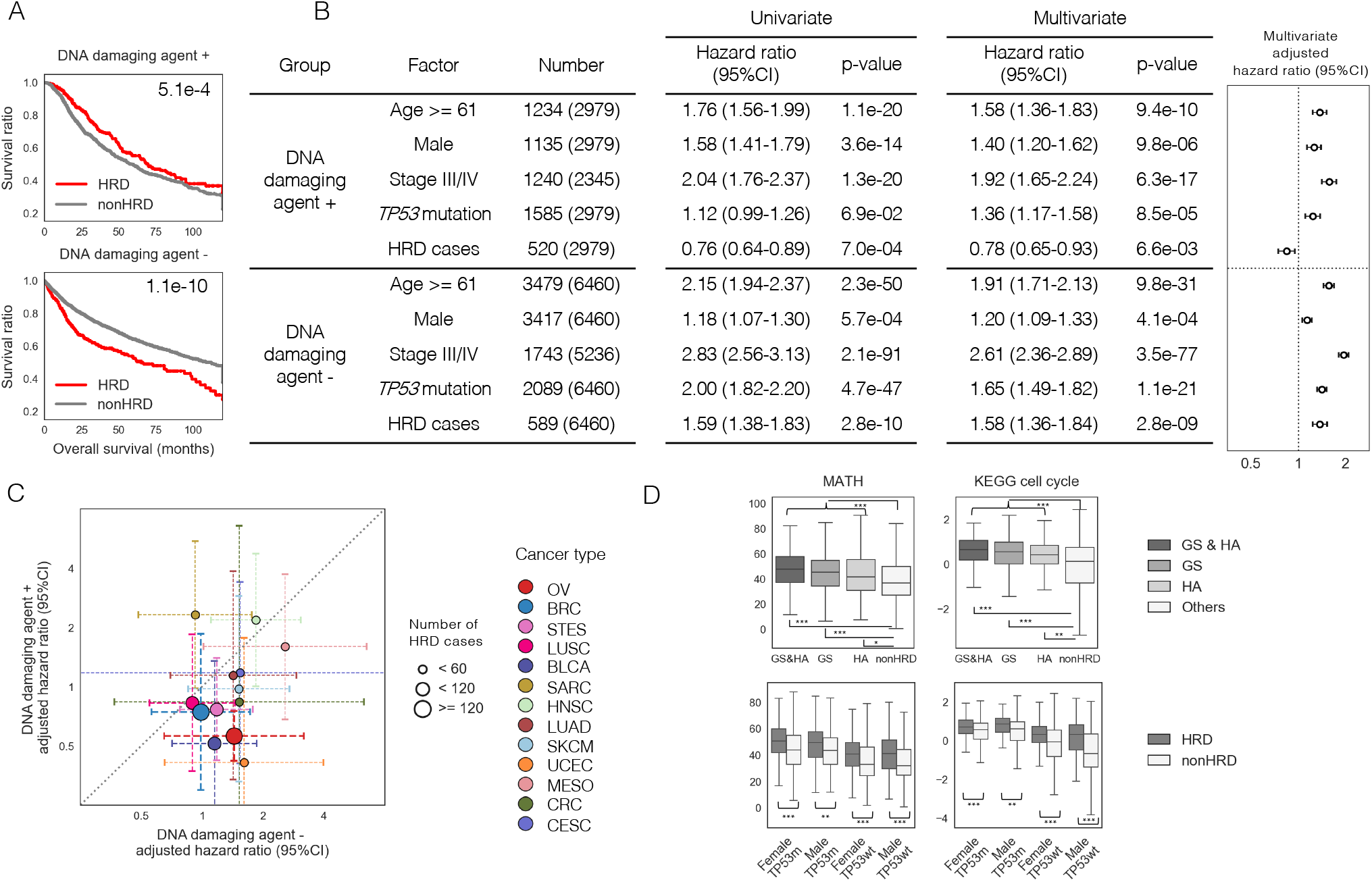
The prognosis of HRD patients differed with and without the administration of DNA-damaging agents. A) Comparison of overall survival between HRD and non-HRD cases by the Kaplan–Meier curve, in two groups divided by the history of DNA-damaging agent use. The number in the upper right corner represents the p-value in the log-rank test. B) Left: univariate and multivariate Cox proportional hazard model analyses with covariates of age, gender, *TP53* mutation, clinical stage, and HRD cases. Right: forest plots showing adjusted hazard ratios with 95%CIs from the multivariate Cox analysis. C) Comparison of the two adjusted hazard ratios of HRD cases for overall survival among 13 cancer types that have five or more comparable cases. As in B, samples were divided into two groups by DNA-damaging agent use and analyzed by multivariate Cox proportional hazard model to calculate the adjusted hazard ratios with 95%CIs. D) Comparisons of MATH index and KEGG cell cycle scores between HRD cases and non-HRD cases, or in the four groups divided by gender and *TP53* mutation. The p-values by the Mann–Whitney U test were 1.1 × 10^−42^ (upper left), 3.2 × 10^−43^ (upper right), and 1.5 × 10^−13^, 7.7 × 10^−7^, 9.3 × 10^−9^ and 4.4 × 10^−14^ (lower left), and 3.6 × 10^−7^, 1.4 × 10^−9^, 4.8 × 10^−9^, and 3.1 × 10^−30^ (lower right), respectively. *, **, and *** stand for P < 0.01, P < 1 × 10^−5^, and P < 1 × 10^−7^ in the Mann–Whitney U test.

Furthermore, in the Cox proportional hazard multivariate analysis with covariates of age, gender, stage, and *TP53* mutation, HRD was a good independent prognostic factor in the DNA-damaging agent group (adjusted hazard ratio 0.78, p=6.6 × 10^−3^), whereas it was a poor independent prognostic factor in the non-DNA-damaging agent group (adjusted hazard ratio 1.58, p=2.8 × 10^−9^) (Fig. 5B). Similar analyses were performed for each of the 13 cancer types that contained five or more comparable cases in both groups on the use of DNA-damaging agents. As a result, in most cancer types, the adjusted hazard ratios of HRD were lower in the DNA-damaging agent group than in the non-DNA-damaging agent group (Fig. 5C, Additional file 2: Table S1). These results suggest that DNA-damaging agents would improve the prognosis of HRD cases, regardless of the cancer type.

Next, we examined why the survival outcome of HRD cases is worse in the absence of DNA-damaging agents than in non-HRD cases. The MATH index^46^, an indicator of intratumor heterogeneity, was higher in HRD cases than in non-HRD cases (Fig. 5D). This result suggests that HRD-induced genomic instability causes increased intratumor heterogeneity and may lead to a poor prognosis. Also, the gene expression-based ssGSEA score of the KEGG cell cycle signature (see Methods) was higher in HRD cases than in non-HRD cases (Fig. 5D). This result suggests that HRD tumors have a high proliferation capacity.

In addition, we performed a similar analysis without the stratification by gender and *TP53* mutation status, using universal cutoff values for HRD score and Sig3 ratio (Additional file 1: Figure S11). Although the associations between gene expression profiles, drug administration and survival outcomes of HRD cases showed similar trends to those with the stratification, the characteristics of the determined HRD cases, such as differences in hazard ratios and p-values in the survival analysis, appeared to be weaker (adjusted hazard ratio: 1.37 vs 0.83, p=7.7× 10^−4^ vs 0.043, respectively, Additional file 1: Figure S11).

### Validation of high sensitivity of DNA-damaging agents to HRD tumors in cell lines

Using datasets of human cancer cell lines from the Cancer Cell Line Encyclopedia (CCLE), we performed exactly the same analysis as described above to reproduce the association between HRD status and chemotherapy sensitivity.

As a result, genomic scar scores were elevated only when HRR pathway gene mutations were involved in the locus-specific LOH (Additional file 1: Figure S12AB), mutations without LOH were enriched in hypermutators without elevation of these scores (Additional file 1: Figure S12C). Stratified analysis by gender origin and presence of TP53 mutation also showed similar trends to the clinical samples; the group of female origin with *TP53* mutations tended to have the highest genomic scar scores, the groups with *TP53* mutations tended to have higher HRD score in the absence of HRR pathway gene alterations (Additional file 1: Figure S13A), and only the group of male origin without *TP53* mutations did not show a positive correlation between HRD score and Sig3 ratio (Additional file 1: Figure S13B, F). We determined the genomic scar high (GS) cell lines with the optimal cutoff values of HRD score and Sig3 ratio calculated from the AUC curve for each of the four groups divided by gender and *TP53* mutation status (Additional file 1: Figure S13B, C, F). For samples with no information on gender or *TP53* mutation, the cutoff values were calculated using data from all samples (Additional file 1: Figure S13D).

After identifying GS and HA samples (Additional file 1: Figure S13E), we examined the IC50 values of 198 compounds for these cell lines using the datasets of the Genomics of Drug Sensitivity in Cancer (GDSC). Consequently, almost all DNA-damaging drugs showed higher sensitivity to HRD tumors than to non-HRD tumors (Additional file 2: Table S5, Fig. 6A). Furthermore, this category of drugs showed higher sensitivity to HRD tumors compared to other types of drugs. (Fig. 6B).

**Fig.6.**
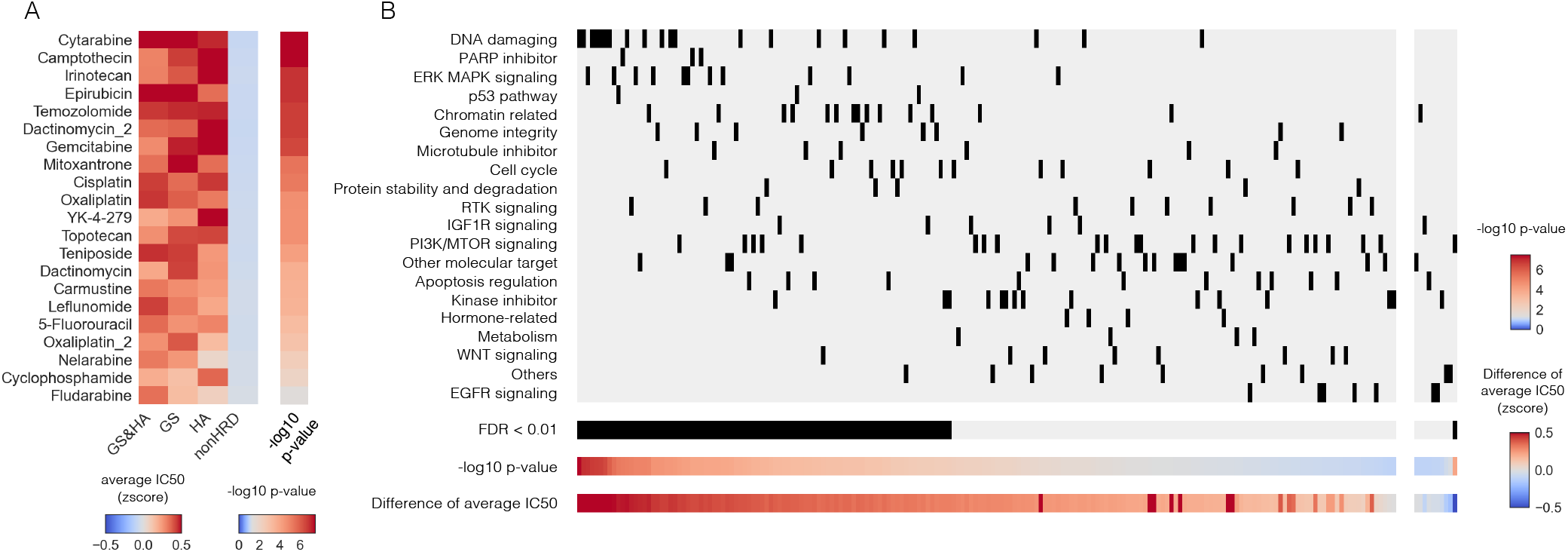
Association between HRD status and chemotherapy sensitivity in CCLE cell lines. A) Most of the DNA-damaging agents showed higher sensitivity in HRD tumors than in non-HRD tumors. The left heatmap indicates the average IC50 for each group. The right heatmap shows p-values of the Mann–Whitney test comparing the IC50 of GS and/or HA samples to non-HRD samples. The center of the color bar is adjusted to P = 0.05. B) Association between drug sensitivity and drug type for cell lines with HRD status. The IC50 values were compared between HRD and non-HRD samples for 198 compounds examined in GDSC2. These compounds were divided into the left or right by positive or negative difference in average IC50, and then arranged in order of decreasing or increasing -log10 p-values on the x-axis. The drug categories were arranged on the Y-axis in order of decreasing mean of -log10 p-value. HRD samples showed significant sensitivity to several drugs (FDR<0.01), especially high sensitivity to DNA-damaging drugs.

## Discussion

Many of the HRR pathway genes are considered to be tumor suppressor genes, which typically require biallelic alterations for actual loss of function^21^. In our study, most gene mutations in the HRR pathway showed increased genomic scar scores only when accompanied by locus-specific LOH (Figs. 1A,1B), whereas mutations without LOH were considered to be passengers (Fig. 1C). These results strongly suggest that to acquire the HRD phenotype, a genetic mutation in the HRR pathway must have the contralateral wild-type allele loss. Furthermore, the frequency of related locus-specific LOH in HRR pathway genes varied widely by gene and cancer type (Fig 2A). These observations are clinically important because they indicate that if a cancer has a genetic mutation in the HRR pathway, its pathogenic importance should be determined by assessing whether it is accompanied by locus-specific LOH, such as by gene panel testing. In addition, we used a combination of FACETS^23^ and ASCAT^22^ to examine the presence of locus-specific LOH and, similarly, used ABSOLUTE^28^ and ASCAT^22^ to test for homozygous deletions. These integrated analyses provided a clearer association between biallelic changes and genomic scar scores than when only a single method was used (Additional file 1: Figure S2, S4), showing that using ensembles of algorithms can improve robustness. While one study reported a benefit of PARP inhibitors in *BRCA*-associated cancers even with monoallelic *BRCA1/2* alterations^25^, another study showed that tumors with germline *BRCA1/2* mutations without locus-specific LOH have a low HRD score and low drug sensitivity^24^. Considering that all of these studies used only one algorithm, the differences in results may be due to difference in methods. Recently, several clinical trials of PARP inhibitors have been conducted in which HRR pathway gene mutations have been examined as biomarkers (e.g., NCT03209401, NCT03377556, NCT04123366). However, few have assessed whether these mutations are biallelic or not. Our data indicate that the clinical relevance of these variants is a very important consideration in future clinical trials and that a robust method for determining the zygosity of these variants needs to be established.

Biallelic *ATM* mutations were the second most common HRR alteration after *BRCA1/2* (Fig. 3B). ATM is well known to have an important function in HRR from the results of many previous studies in Ataxia-telangiectasia. Recent studies have shown that *ATM*-deficient cells were sensitive to PARP inhibitors^47,48^, similarly that low *ATM* gene expression was associated with sensitivity to PARP inhibitors in some clinical trials^49,50^. We found that biallelic *ATM* and *TP53* mutations were significantly mutually exclusive (Fig. 4A). The mutual exclusivity of these two has also been reported in hematologic^51,52^ and breast^53^ cancers. Since *ATM* has been reported to activate *TP53* through the degradation of *MDM2* after sensing DSBs and induce tumor suppressive effects including cell cycle delay, arrest, and apoptosis, this mutual exclusivity is plausibly due to the similarities between *ATM* and *TP53* mutations in terms of disrupting the cell cycle checkpoint mechanism for cancer cells. Also, the relatively lower HRD scores in *ATM*-mutated cases (Additional file 1: Figure S3) may be explained by this mutual exclusivity.

In our analysis, HRD score was higher in tumors with *TP53* mutations (Fig. 2C). From previous studies on *TP53* mutations and HRD status, some reported that *TP53* mutations activate the HRR pathway under certain circumstances^54^, while others suggest that they did not directly associate with HRD^55,56^. In our data, *TP53* mutation did not correlate with the Sig3 ratio (Fig. 2C), while *TP53* mutated cases, even without HRR alteration, had higher HRD score than *TP53* wild-type cases (Fig. 3A). In a very recent study of prostate cancer, *TP53* mutations were reported to be associated with increased HRD scores independently of HRR pathway gene alterations^57^. These observations suggest that the high HRD score in *TP53* mutated cases is not necessarily due to true HRD but, rather, due to chromosomal instability caused by mechanisms other than HRD (e.g., loss of cell-cycle checkpoint mechanisms).

We observed that not only the two genomic scar scores themselves, but also their correlations, differed greatly depending on gender and *TP53* mutation status (Fig. 3). Specifically, the correlation between HRD score and Sig3 ratio was the strongest in women with *TP53* mutations, though weaker in the other groups (Fig. 3B). Also, the AUC values to determine HRR pathway gene-altered cases were lower in the groups other than women with *TP53* mutations (Fig. 3C). Although the analysis using a pair of universal cutoffs without stratification by gender or *TP53* mutation was still able to extract cases with HRD characteristics (Additional file 1: Figure S11), these cutoff values were impacted, resulting in over-estimation in women with *TP53* mutations and under-estimation in the other groups, thus weakening the characteristics of the determined HRD cases (Additional file 1: Figure S11). The HRD score and Sig3 ratio were originally developed in a rather limited analysis to identify *BRCA1/2* mutant tumors in breast and ovarian cancers^14-16,18,19^ and were reported to have low correlation^19^. Based on the findings after stratification, we propose that differences in gender and *TP53* should be taken into account when using these genomic scores to determine HRD cases. For the groups other than women with *TP53* mutations, a more appropriate measure other than the HRD/Sig3 scores could further strengthen analysis of HRD. Given that large-scale multi-omics databases are being constructed worldwide and that new methods for evaluating HRD using whole-genome sequencing are emerging^58,59^, there is opportunity to develop more robust methods for identifying HRD cases in pan-cancer contexts.

One of the major differences between the cells of men and women is the presence of X-chromosome inactivation. And the loss of X-chromosome inactivation in some female cancers has long been known as the loss of the Barr body^60^. In the present pan-tumor analysis, we found that the loss of X-chromosome inactivation was associated with female HRD tumor cells (Additional file 1: Figure S10). This suggests that gender differences in tumor HRD status and genomic scar scores are related to X-chromosome inactivation (Fig. 3A). There have been no reports on the association between the genomic scar-based HRD status and the X chromosome, and more detailed studies are needed in the future.

HRR pathway gene alterations were detected in only half of the genomic scar high (GS) cases (Fig. 4A). There are many possible causative factors for HRD other than those analyzed in this study. For instance, abnormalities in histone modifications and chromatin remodeling that operate in relation to DSB repair are reported to be strongly associated with HRD^61^. More recently, oncometabolites have been reported to result in HRD in cancer cells^62,63^. Thus, mechanisms that have not yet been fully elucidated may be involved in HRD, and further studies are warranted.

Recently, an emerging therapeutic strategy for cancer is the idea that cancers with defects in a certain DNA damage repair mechanism should be vulnerable to drugs that are targeting the same mechanism^44^. In this context, HRD tumors would be susceptible to drugs that induce DNA damage for which HRR is essential. Platinum compounds, alkylating agents, and some antibiotic-derived anticancer drugs (e.g., mitomycin C) cause DNA damage that requires HRR for recovery by forming inter- and intra-strand crosslinks between DNAs^64^. In practice, the efficacy of platinum agents for HRD tumors is well recognized in ovarian and breast cancers^1^. Additionally, topoisomerase inhibitors and anthracycline anti-tumor antibiotics form covalent enzyme-DNA cleavage complexes that block the relegation of cleaved DNA ends, resulting in DNA strand breaks and inhibition of DNA synthesis^45^. Also, antimetabolites deplete the pool of available deoxyribonucleoside triphosphates by inhibiting enzymes necessary for nucleic acid synthesis, or they mimic natural substrates and are incorporated into nascent DNA strands to inhibit DNA synthesis^45^. In other words, all of these drugs hinder DNA replication and stall the replication fork, causing replication stress^45^. Since these stalled forks can only be repaired through HRR, replication stress will accumulate in HRR-dysfunctional cells up to a critical level after which replication forks collapse, leading to DNA double-strand breaks and cell death^45^. Therefore, in this study, we included drugs with these mechanisms as the DNA-damaging agents and found high sensitivity to HRD tumors in both clinical samples and cell lines.

In general, the standard of care for cancer treatment is selected on the basis of organ-specific clinicopathological factors derived from the results of the many randomized trials that have been conducted up to that time. However, our study demonstrated the potential benefit of DNA-damaging drug administration for HRD tumors as defined by pan-cancer genomic analysis (Fig. 5). The results indicate that treatment personalization beyond cancer type based on molecular phenotype including HRD status has substantial potential. Since the number of cases treated with PARP inhibitors in the TCGA cohort is highly limited, further investigations and publicly-accessible datasets are needed to determine if pan-cancer HRD analysis could predict sensitivity to PARP inhibitors.

## Conclusion

This comprehensive pan-solid cancer HRD analysis revealed that stratified analysis by gender and *TP53* mutation status can more accurately assess HRD status, and that DNA-damaging drugs could be beneficial for HRD cases across cancer types, suggesting that HRD is useful as a tumor-agnostic therapeutic biomarker. Based on the results, it is rational to expect improvements in the implementation of personalized cancer medicine based on HRD. In parallel, further diagnostic methods can be developed for the pinpoint identification of clinically significant HRD cases.

## Methods

### Patient selection and clinical data

Among the 10,967 cases integrated into the TCGA Pan-Cancer Atlas at cBioPortal, the cases with acute myeloid leukemia, diffuse large B-cell lymphoma, and thymoma, and the cases without the MC3 somatic mutation profiles^65^ were excluded. The remaining 9,847 cases were analyzed (Additional file 2: Table S2).

Clinical data were obtained from cBioPortal and Broad GDAC. Regarding the drugs administered in each case, we first reviewed and manually unified treatment annotations (e.g., for drug-spelling errors), then subsequently extracted individual drugs when they were part of combination therapies. Next, we separated the anti-tumor agents into the following categories: alkylating agents, antibiotics, antimetabolites, platinum, topoisomerase, microtubule inhibitors, molecular-target agents, immune checkpoint inhibitors, others, and those unresolvable from annotations. Finally, the drugs in the first five categories were defined as DNA-damaging drugs (Additional file 2: Table S3). Fewer than 10 cases were treated with PARP inhibitors, all priorly treated by DNA-damaging drugs.

### Genomic scar scores

As an indicator of HRD based on chromosomal structural changes, LST, TAI, and LOH scores published in Pan-Cancer Atlas studies were obtained from the Genomic Data Commons (GDC) website. The sum of these three scores was used as the HRD score. In addition, as a score of HRD based on the pattern of single base substitutions, the mutational signature 3 value was calculated using the method previously described by Jonsson et al.^25^. To be more specific, we obtained somatic mutation profiles published in the MC3 project^65^ from the above-mentioned GDC website and, using 30 cancer mutational signatures from COSMIC version 2 (https://cancer.sanger.ac.uk/cosmic/signatures_v2) as a reference, calculated the contribution ratio to signature 3 as the “Sig3 ratio” by applying non-negative matrix factorization.

### Definition of homologous recombination repair (HRR) pathway genes

Based on the literature^66^ we selected *BRCA1, BRCA2, ATM, ATR, BARD1, BLM, BRIP1, CDK12, CHEK1, CHEK2, FANCA, FANCC, FANCD2, FANCE, FANCF, FANCI, FANCL, FANCM, MRE11, NBN, PALB2, RAD50, RAD51, RAD51B, RAD51C, RAD51D, RAD52, RAD54L*, and *RPA1* as HRR pathway genes (n=29).

For germline gene mutations, we obtained the annotation information from the supplement data of the previous paper, in which 10,339 cases from TCGA were examined for pathological germline variants^67^ available on the GDC website. From the data, we extracted 412 cases who had germline mutations in the above HRR pathway genes with annotation of “likely pathogenic” or “pathogenic” in “Overall Classification” column.

For somatic gene mutations, we extracted the annotation information from cBioPortal except for both mutations and copy number alterations of unknown significance.

### Locus-specific LOH status

The raw SNP genotyping array data of normal and tumor pairs were obtained from the GDC legacy archive, and the segmented genome-wide allele-specific copy number profiles were calculated using PennCNV^68^ and ASCAT^22^ The whole-exome sequencing data of normal and tumor pairs were obtained from the GDC data portal, and the allele-specific copy number was calculated using FACETS^23^ We checked the estimated copy numbers of the minor allele at the segment located in the locus of each of the above germline and somatic mutations; when the copy number of the minor allele was equal to zero, it was determined to be “LOH,” and when it was one or more, it was determined to be “non-LOH.” When it was unavailable, it was determined to be “unknown.” Then, we examined the concordance between the results from ASCAT and FACET for the presence of locus-specific LOH in each variant (Additional file 1: Figure S2). The variants that were determined to have LOH by both algorithms, or to have LOH in one algorithm and to be unknown in the other, showed higher genomic scar scores than the other classification outcome groups. From these observations, we determined these variants as those accompanying locus-specific LOH (Additional file 1: Figure S2).

### Homozygous deletion status

From the allele-specific copy number profiles calculated by ASCAT^22^ as described above, segmented regions with a total copy number equal to zero were extracted as homozygously deleted regions in each sample. Additionally, we obtained segmented copy number data calculated by ABSOLUTE^28^ and published in the Pan-Cancer Atlas studies from the GDC website and extracted the regions annotated as “Homozygous_deletion” per sample. Using each annotation separately, for each HRR pathway gene in each case, a gene was determined to have homozygous deletion when part or all of the locus was contained in the above homozygously deleted regions. The results calculated from ASCAT and ABSOLUTE were combined to form the final annotation (Additional file 1: Figure S4A).

### DNA promoter methylation

In a previous report, Knijnenburg et al. comprehensively investigated all of the TCGA samples for DNA promoter methylation status in 276 genes involved in DNA-damage repair and determined cases with methylation-driven gene silencing in stringent criteria^35^. From their published data, we found one or more methylated sample in *BRCA1, RAD51C*, and *FANCC2* among the above HRR pathway genes (Additional file 1: Figure S5).

To quantify DNA methylation across the entire X chromosome, DNA promoter methylation data from the Illumina Infinium array published in The Pan-Cancer Atlas Studies were obtained from the GDC website. Among the 20601 probes, the average beta value of 856 probes on the X chromosome was calculated as the amount of X chromosome global methylation for each sample.

### Tumor mutational burden, MSI-high annotation, somatic *POLE*, and *TP53* mutation

From the somatic mutation profile published by the MC3 project^65^, the total number of non-synonymous mutations in each sample was counted and defined as tumor mutational burden (TMB).

Among the patients’ clinical data obtained from cBioPortal, annotations of tumor subtypes including MSI status exist for the TCGA-COAD, READ, UCEC, STAD, and ESCA cohorts. We recorded those cases as MSI-high for further analysis.

We also extracted somatic *POLE* and *TP53* mutation annotation information from cBioPortal, discarding annotations marked as mutations and copy number alterations of unknown significance.

### Gene expression analysis

From the GDC website, we retrieved the published batch effect-corrected mRNA gene expression data published in Pan-Cancer Atlas studies. Expression values of genes whose missing data exceeded 25% of all cases were excluded from the analysis. Otherwise, the missing data were completed with the median value in all other cases as the representative value.

In the original work^38^, the Recombination Proficiency Score (RPS) was calculated as the sum of the microarray-based gene expression values of the Rif1, PARI, Ku80, and RAD51 genes after log2-transformation and median normalization, and being multiplied by −1 for each gene. We modified this method to simply sum up the RNA-based gene expression values of the same four genes after log2-transforming and z-scaling in each gene and named the value reversed RPS (rRPS) score as an indicator of HRD.

The configuration of the TCGA-OV derived HRD signature was as follows. First, we selected 243 ovarian cancer cases for which the Sig3 ratio, HRD score, and mRNA gene expression data were all available. Next, we selected cases that were in the top 50% of both the Sig3 ratio and HRD score, and cases that were in the bottom 50% of both. Between those two groups, differentially expressed genes were analyzed by limma+voom^69^. Finally, we extracted the 76 genes highly expressed in the top group according to certain criteria (logFC > 0.5, AveExpr > −2, adj.P.Val < 0.01) and designated them as TCGA-OV derived HRD signatures (Additional file 2: Table S4). The enrichment score of the gene signatures for each sample was calculated using the ssGSEA algorithm^39^ and was designated as the OV-GS score. Because ovarian cancer was used as a training set, we assigned the OV-GS score values to cancer types other than ovarian cancer.

In addition, the gene sets of KEGG_HOMOLOGOUS_RECOMBINATION (hsa03440, 41 genes) and KEGG_CELL_CYCLE (hsa04110, 124 genes) were obtained from MsigDB (http://www.gsea-msigdb.org/gsea/msigdb/). The only gene found in common in the two KEGG gene sets was *ATM*. These signatures were also used for scoring by ssGSEA and designated as the KEGG-score and the KEGG cell cycle score, respectively.

To assess overall gene expression of the X chromosome, a list of 1666 gene names on the X chromosome was obtained from Ensemble BioMart Release 102 (GRCh38.p13). The enrichment score for each sample was calculated using ssGSEA and named as X score based on this gene set. We also obtained a list of 50 X-chromosome inactivation escaping genes from a previous report^7*0*^ and calculated the X-chromosome inactivation score (XCI-esc score) per sample using ssGSEA.

### MATH score

The MATH score for each sample was calculated from the somatic mutation profile published in the MC3 project^65^, by using the method described in the previous reports^46^.

### Retrieving validation datasets

From the previous literature^25^ and the database of AACR Project GENIE (https://www.aacr.org/professionals/research/aacr-project-genie/), the clinical data, signature 3 values, HRD scores (sum of the LOH, LST, and TAI scores), and somatic mutation profiles from the 815 samples of 794 multiple cancer cases were obtained. Of these, two samples from two cases with hematological and lymphatic cancers were excluded from analysis.

### Data analysis of CCLE cell lines

We obtained the gene mutation data from the DepMap portal (https://depmap.org/portal/) and determined that mutations marked as “damaging” in the “variant annotation” were pathogenic. The total number of nonsynonymous mutations in each cell line was defined as TMB. We obtained allele-specific copy number data analyzed by ABSOLUTE^28^ from the same data source and determined whether the above individual pathogenic mutations were accompanied by a locus-specific LOH: with LOH if the number of minor alleles at the locus was equal to 0, without LOH if the number was greater than one, and unknown if the number was unknown. For *BRCA1* methylation, RNAseq gene expression and DNA methylation data were obtained, and samples with promoter methylation beta values of 0.3 or higher and gene expression below the median of all samples were determined to have methylation-associated silencing (Additional file 1: Figure S12D). HRD scores were calculated using scarHRD^71^ with the allele-specific copy number and average ploidy data available in the COSMIC cell line project (https://cancer.sanger.ac.uk/cell_lines). Based on copy number aberration analysis for each gene in the same project, genes with a total copy number of zero were determined to have homozygous deletions. The single-base-substitution pattern in the 96 trinucleotide contexts for each cell line was obtained from the published data of Petljak et al.^72^ and the Sig3 ratio was calculated using the same method as described above ^25^. The inferred MSI status was obtained from the published data of Ghandi et al.^73^. IC50 data of 198 compounds for each sample were obtained from Genomics of Drug Sensitivity in Cancer (GDSC, https://www.cancerrxgene.org/) released as version 2. These compounds were classified into the following 20 categories based on the information of target molecules and target pathways (Additional file 2: Table S5): Apoptosis regulation, Cell cycle, Chromatin related, DNA damaging, EGFR signaling, ERK MAPK signaling, Genome integrity, Hormone-related, IGF1R signaling, Kinase inhibitor, Metabolism, Microtubule inhibitor, Other molecular target, Others, PARP inhibitor, PI3K/MTOR signaling, Protein stability and degradation, RTK signaling, WNT signaling, p53 pathway.

### Statistical analyses

All statistical analyses in this study were performed in Python (3.8.6). The Mann–Whitney U test, chi-square test, and Spearman’s rank correlation coefficient test were performed using SciPy (1.6.0). Survival analyses including the Kaplan–Meier curve, log-rank test, and Cox proportional hazard regression model were performed using Lifelines (0.25.8). Unless otherwise noted, a p-value < 0.05 was considered statistically significant.

### Availability of data and materials

Open access data and processed data for TCGA samples are available from the cBioPortal (https://www.cbioportal.org/), the Broad GDAC (https://gdac.broadinstitute.org/), and the GDC Pan-Cancer Atlas studies (https://gdc.cancer.gov/about-data/publications/pancanatlas). Controlled access data including germline variant annotations, raw sequence data, and raw SNP array data are available through the dbGaP-authorized access system (study accession phs000178). CCLE cell line datasets and their drug sensitivity data are available in the DepMap portal (https://depmap.org/portal/), the COSMIC cell line project (https://cancer.sanger.ac.uk/cell_lines), and Genomics of Drug Sensitivity in Cancer (https://www.cancerrxgene.org/). The processed data and codes to reproduce the main results of this work are available on the GitHub page (https://github.com/shirotak/pancancer_hrd_analysis). Other codes for preprocessing public or restricted-access data are available from the corresponding author upon reasonable request.

## Additional files

**Additional file 1: Figure S1**. Correlations among LOH, TAI, LST, and HRD scores in all cases. **Figure S2**. Assessment of the locus-specific LOH. **Figure S3**. Distribution of germline and somatic mutations with or without locus-specific LOH in HRR pathway genes excluding *BRCA1/2*. **Figure S4**. Determination of homozygous deletion. **Figure S5**. Correlation between promoter methylation-driven gene silencing of HRR pathway genes and genomic scar signatures. **Figure S6**. Differences in genomic scar scores by gender and *TP53* mutation in non-*BRCA*-associated cancers. **Figure S7**. Cancer types and the number of cases assigned to GS cases and/or HA cases in the four groups divided by gender and the presence of *TP53* mutation. **Figure S8**. Identification of TCGA OV-derived HRD signature. **Figure S9**. Correlations among rRPS score, OV-GS score, and KEGG score. **Figure S10**. Differences in X chromosome inactivation by gender and HRD status. **Figure S11**. Reanalysis of HRD cases without patient stratification by gender and *TP53* mutation. **Figure S12**. Association between HRR pathway gene alterations and genomic scar scores or tumor mutation burden in CCLE cell lines. **Figure S13**. Identification of genomic scar high (GS) cases in CCLE cell lines.

**Additional file 2: Table S1**. Multivariate Cox proportional hazard analysis in the two groups with or without DNA-damaging drug use. **Table S2**. Number of cases analysed in TCGA by cancer type. **Table S3**. Drug administration information for each case. **Table S4**. TCGA-OV derived HRD signature. **Table S5**. Categorized drug information of 198 compounds in GDSC2.

## Authors’ contributions

ST, JBB, and NM conceived the study, performed analyses, wrote/edited the paper. KY, JH, KY, HT, and TK edited the paper and provided discussion. MN, MS, and MM supervised the study. All authors proofread, made comments, and approved the paper.

## References

1. Lord CJ et al. BRCAness revisited. Nat Rev Cancer. 2016 Feb;16(2):110–20. doi: 10.1038/nrc.2015.21. Epub 2016 Jan 18.

2. Dullens B et al. Cancer Surveillance in Healthy Carriers of Germline Pathogenic Variants in BRCA1/2: A Review of Secondary Prevention Guidelines. J Oncol. 2020 Jun 20;2020:9873954. doi: 10.1155/2020/9873954. eCollection 2020.

3. Farmer H et al. Targeting the DNA repair defect in BRCA mutant cells as a therapeutic strategy. Nature. 2005 Apr 14;434(7035):917–21. doi: 10.1038/nature03445.

4. Bryant HE et al. Specific killing of BRCA2-deficient tumours with inhibitors of poly(ADP-ribose) polymerase. Nature. 2005 Apr 14;434(7035):913–7. doi: 10.1038/nature03443.

5. Patel M et al. The role of poly(ADP-ribose) polymerase inhibitors in the treatment of cancer and methods to overcome resistance: a review. Cell Biosci. 2020 Mar 11;10:35. doi: 10.1186/s13578-020-00390-7. eCollection 2020.

6. Nizialek E et al. PARP Inhibitors in Metastatic Prostate Cancer: Evidence to Date. Cancer Manag Res. 2020 Sep 7;12:8105–8114. doi: 10.2147/CMAR.S227033. eCollection 2020.

7. Póti Á et al. Correlation of homologous recombination deficiency induced mutational signatures with sensitivity to PARP inhibitors and cytotoxic agents. Genome Biol. 2019 Nov 14;20(1):240. doi: 10.1186/s13059-019-1867-0.

8. Stover EH et al. Clinical assays for assessment of homologous recombination DNA repair deficiency. Gynecol Oncol. 2020 Dec;159(3):887–898. doi: 10.1016/j.ygyno.2020.09.029. Epub 2020 Oct 2.

9. Sandhu SK et al. The poly(ADP-ribose) polymerase inhibitor niraparib (MK4827) in BRCA mutation carriers and patients with sporadic cancer: a phase 1 dose-escalation trial. Lancet Oncol. 2013 Aug;14(9):882–92. doi: 10.1016/S1470-2045(13)70240-7. Epub 2013 Jun 28.

10. de Bono J et al. Phase I, Dose-Escalation, Two-Part Trial of the PARP Inhibitor Talazoparib in Patients with Advanced Germline BRCA1/2 Mutations and Selected Sporadic Cancers. Cancer Discov. 2017 Jun;7(6):620–629. doi: 10.1158/2159-8290.CD-16-1250. Epub 2017 Feb 27.

11. A phase II study of talazoparib (BMN 673) in patients with homologous recombination repair deficiency (HRRD) positive stage IV squamous cell lung cancer (Lung-MAP Sub-Study, S1400G). Owonikoko TK et al. JCO 37, 9022–9022 (2019). doi: 10.1200/JCO.2019.37.15_suppl.9022

12. Hutter C et al. The Cancer Genome Atlas: Creating Lasting Value beyond Its Data. Cell. 2018 Apr 5;173(2):283–285. doi: 10.1016/j.cell.2018.03.042.

13. International Cancer Genome Consortium et al. International network of cancer genome projects. Nature. 2010 Apr 15;464(7291):993–8. doi: 10.1038/nature08987.

14. Birkbak NJ et al. Telomeric allelic imbalance indicates defective DNA repair and sensitivity to DNA-damaging agents. Cancer Discov. 2012 Apr;2(4):366–375. doi: 10.1158/2159-8290.CD-11-0206. Epub 2012 Mar 22.

15. Popova T et al. Ploidy and large-scale genomic instability consistently identify basal-like breast carcinomas with BRCA1/2 inactivation. Cancer Res. 2012 Nov 1;72(21):5454–62. doi: 10.1158/0008-5472.CAN-12-1470. Epub 2012 Aug 29.

16. Abkevich V et al. Patterns of genomic loss of heterozygosity predict homologous recombination repair defects in epithelial ovarian cancer. Br J Cancer. 2012 Nov 6;107(10):1776–82. doi: 10.1038/bjc.2012.451. Epub 2012 Oct 9.

17. Telli ML et al. Homologous Recombination Deficiency (HRD) Score Predicts Response to Platinum-Containing Neoadjuvant Chemotherapy in Patients with Triple-Negative Breast Cancer. Clin Cancer Res. 2016 Aug 1;22(15):3764–73. doi: 10.1158/1078-0432.CCR-15-2477. Epub 2016 Mar 8.

18. Alexandrov LB et al. Signatures of mutational processes in human cancer. Nature. 2013 Aug 22;500(7463):415–21. doi: 10.1038/nature12477. Epub 2013 Aug 14.

19. Polak P et al. A mutational signature reveals alterations underlying deficient homologous recombination repair in breast cancer. Nat Genet. 2017 Oct;49(10):1476–1486. doi: 10.1038/ng.3934. Epub 2017 Aug 21.

20. Gou R et al. Application and reflection of genomic scar assays in evaluating the efficacy of platinum salts and PARP inhibitors in cancer therapy. Life Sci. 2020 Nov 15;261:118434. doi: 10.1016/j.lfs.2020.118434. Epub 2020 Sep 14.

21. Knudson AG et al. Two genetic hits (more or less) to cancer. Nat Rev Cancer. 2001 Nov;1(2):157–62. doi: 10.1038/35101031.

22. Van Loo P et al. Allele-specific copy number analysis of tumors. Proc Natl Acad Sci U S A. 2010 Sep 28;107(39):16910–5. doi: 10.1073/pnas.1009843107. Epub 2010 Sep 13.

23. Shen R et al. FACETS: allele-specific copy number and clonal heterogeneity analysis tool for high-throughput DNA sequencing. Nucleic Acids Res. 2016 Sep 19;44(16):e131. doi: 10.1093/nar/gkw520. Epub 2016 Jun 7.

24. Maxwell KN et al. BRCA locus-specific loss of heterozygosity in germline BRCA1 and BRCA2 carriers. Nat Commun. 2017 Aug 22;8(1):319. doi: 10.1038/s41467-017-00388-9.

25. Jonsson P et al. Tumour lineage shapes BRCA-mediated phenotypes. Nature. 2019 Jul;571(7766):576–579. doi: 10.1038/s41586-019-1382-1. Epub 2019 Jul 10.

26. Birkbak NJ et al. Tumor mutation burden forecasts outcome in ovarian cancer with BRCA1 or BRCA2 mutations. PLoS One. 2013 Nov 12;8(11):e80023. doi: 10.1371/journal.pone.0080023. eCollection 2013.

27. Pellegrino B et al. Homologous Recombination Repair Deficiency and the Immune Response in Breast Cancer: A Literature Review. Transl Oncol. 2020 Feb;13(2):410–422. doi: 10.1016/j.tranon.2019.10.010. Epub 2020 Jan 2.

28. Carter SL et al. Absolute quantification of somatic DNA alterations in human cancer. Nat Biotechnol. 2012 May;30(5):413–21. doi: 10.1038/nbt.2203.

29. Grundy MK et al. Regulation and pharmacological targeting of RAD51 in cancer. NAR Cancer. 2020 Sep;2(3):zcaa024. doi: 10.1093/narcan/zcaa024. Epub 2020 Sep 25.

30. Sokol ES et al. Pan-Cancer Analysis of BRCA1 and BRCA2 Genomic Alterations and Their Association With Genomic Instability as Measured by Genome-Wide Loss of Heterozygosity. JCO Precis Oncol. 2020;4:442–465. doi: 10.1200/po.19.00345. Epub 2020 Apr 30.

31. Shen H et al. Integrated Molecular Characterization of Testicular Germ Cell Tumors. Cell Rep. 2018 Jun 12;23(11):3392–3406. doi: 10.1016/j.celrep.2018.05.039.

32. Cavallo F et al. Reduced proficiency in homologous recombination underlies the high sensitivity of embryonal carcinoma testicular germ cell tumors to Cisplatin and poly (adp-ribose) polymerase inhibition. PLoS One. 2012;7(12):e51563. doi: 10.1371/journal.pone.0051563. Epub 2012 Dec 12.

33. Cavallo F et al. Assessing Homologous Recombination and Interstrand Cross-Link Repair in Embryonal Carcinoma Testicular Germ Cell Tumor Cell Lines. Methods Mol Biol. 2021;2195:113–123. doi: 10.1007/978-1-0716-0860-9_9.

34. Donehower LA et al. Integrated Analysis of TP53 Gene and Pathway Alterations in The Cancer Genome Atlas. Cell Rep. 2019 Jul 30;28(5):1370-1384.e5. doi: 10.1016/j.celrep.2019.07.001.

35. Knijnenburg TA et al. Genomic and Molecular Landscape of DNA Damage Repair Deficiency across The Cancer Genome Atlas. Cell Rep. 2018 Apr 3;23(1):239-254.e6. doi: 10.1016/j.celrep.2018.03.076.

36. Agnese DM et al. Battle of the BRCA1/BRCA2 (offspring) sex ratios: truth or consequences. J Med Genet. 2006 Mar;43(3):201–2. doi: 10.1136/jmg.2004.028977. Epub 2005 Jul 20.

37. Moynahan ME et al. The cancer connection: BRCA1 and BRCA2 tumor suppression in mice and humans. Oncogene. 2002 Dec 16;21(58):8994–9007. doi: 10.1038/sj.onc.1206177.

38. Pitroda SP et al. DNA repair pathway gene expression score correlates with repair proficiency and tumor sensitivity to chemotherapy. Sci Transl Med. 2014 Mar 26;6(229):229ra42. doi: 10.1126/scitranslmed.3008291.

39. Barbie DA et al. Systematic RNA interference reveals that oncogenic KRAS-driven cancers require TBK1. Nature. 2009 Nov 5;462(7269):108–12. doi: 10.1038/nature08460. Epub 2009 Oct 21.

40. Kang J et al. A DNA repair pathway-focused score for prediction of outcomes in ovarian cancer treated with platinum-based chemotherapy. J Natl Cancer Inst. 2012 May 2;104(9):670–81. doi: 10.1093/jnci/djs177. Epub 2012 Apr 13.

41. Ouyang Y et al. Inhibition of Atm and/or Atr disrupts gene silencing on the inactive X chromosome. Biochem Biophys Res Commun. 2005 Nov 25;337(3):875–80. doi: 10.1016/j.bbrc.2005.09.122. Epub 2005 Sep 29.

42. ElInati E et al. DNA damage response protein TOPBP1 regulates X chromosome silencing in the mammalian germ line. Proc Natl Acad Sci U S A. 2017 Nov 21;114(47):12536–12541. doi: 10.1073/pnas.1712530114. Epub 2017 Nov 7.

43. Vančevska A et al. SMCHD1 promotes ATM-dependent DNA damage signaling and repair of uncapped telomeres. EMBO J. 2020 Apr 1;39(7):e102668. doi: 10.15252/embj.2019102668. Epub 2020 Feb 21.

44. Pilié PG et al. State-of-the-art strategies for targeting the DNA damage response in cancer. Nat Rev Clin Oncol. 2019 Feb;16(2):81–104. doi: 10.1038/s41571-018-0114-z.

45. Dobbelstein M et al. Exploiting replicative stress to treat cancer. Nat Rev Drug Discov. 2015 Jun;14(6):405–23. doi: 10.1038/nrd4553. Epub 2015 May 8.

46. Mroz EA et al. MATH, a novel measure of intratumor genetic heterogeneity, is high in poor-outcome classes of head and neck squamous cell carcinoma. Oral Oncol. 2013 Mar;49(3):211–5. doi: 10.1016/j.oraloncology.2012.09.007. Epub 2012 Oct 15.

47. Schmitt A et al. ATM Deficiency Is Associated with Sensitivity to PARP1-and ATR Inhibitors in Lung Adenocarcinoma. Cancer Res. 2017 Jun 1;77(11):3040–3056. doi: 10.1158/0008-5472.CAN-16-3398. Epub 2017 Mar 31.

48. Perkhofer L et al. ATM Deficiency Generating Genomic Instability Sensitizes Pancreatic Ductal Adenocarcinoma Cells to Therapy-Induced DNA Damage. Cancer Res. 2017 Oct 15;77(20):5576–5590. doi: 10.1158/0008-5472.CAN-17-0634. Epub 2017 Aug 8.

49. Bang YJ et al. Randomized, Double-Blind Phase II Trial With Prospective Classification by ATM Protein Level to Evaluate the Efficacy and Tolerability of Olaparib Plus Paclitaxel in Patients With Recurrent or Metastatic Gastric Cancer. J Clin Oncol. 2015 Nov 20;33(33):3858–65. doi: 10.1200/JCO.2014.60.0320. Epub 2015 Aug 17.

50. Bang YJ et al. Olaparib in combination with paclitaxel in patients with advanced gastric cancer who have progressed following first-line therapy (GOLD): a double-blind, randomised, placebo-controlled, phase 3 trial. Lancet Oncol. 2017 Dec;18(12):1637–1651. doi: 10.1016/S1470-2045(17)30682-4. Epub 2017 Nov 2.

51. Pettitt AR et al. p53 dysfunction in B-cell chronic lymphocytic leukemia: inactivation of ATM as an alternative to TP53 mutation. Blood. 2001 Aug 1;98(3):814–22. doi: 10.1182/blood.v98.3.814.

52. Greiner TC et al. Mutation and genomic deletion status of ataxia telangiectasia mutated (ATM) and p53 confer specific gene expression profiles in mantle cell lymphoma. Proc Natl Acad Sci U S A. 2006 Feb 14;103(7):2352–7. doi: 10.1073/pnas.0510441103. Epub 2006 Feb 3.

53. Weigelt B et al. The Landscape of Somatic Genetic Alterations in Breast Cancers From ATM Germline Mutation Carriers. J Natl Cancer Inst. 2018 Sep 1;110(9):1030–1034. doi: 10.1093/jnci/djy028.

54. Moureau S et al. A role for the p53 tumour suppressor in regulating the balance between homologous recombination and non-homologous end joining. Open Biol. 2016 Sep;6(9):160225. doi: 10.1098/rsob.160225.

55. Willers H et al. Dissociation of p53-mediated suppression of homologous recombination from G1/S cell cycle checkpoint control. Oncogene. 2000 Feb 3;19(5):632–9. doi: 10.1038/sj.onc.1203142.

56. Boehden GS et al. p53 mutated in the transactivation domain retains regulatory functions in homology-directed double-strand break repair. Oncogene. 2003 Jun 26;22(26):4111–7. doi: 10.1038/sj.onc.1206632.

57. Lotan TL et al. Homologous recombination deficiency (HRD) score in germline BRCA2-versus ATM-altered prostate cancer. Mod Pathol. 2021 Jan 18. doi: 10.1038/s41379-020-00731-4.

58. Davies H et al. HRDetect is a predictor of BRCA1 and BRCA2 deficiency based on mutational signatures. Nat Med. 2017 Apr;23(4):517–525. doi: 10.1038/nm.4292. Epub 2017 Mar 13.

59. Nguyen L et al. Pan-cancer landscape of homologous recombination deficiency. Nat Commun. 2020 Nov 4;11(1):5584. doi: 10.1038/s41467-020-19406-4.

60. Barr ML et al. Chromosomes, sex chromatin, and cancer. Proc Can Cancer Conf. 1957;2:3–16.

61. Aleksandrov R et al. The Chromatin Response to Double-Strand DNA Breaks and Their Repair. Cells. 2020 Aug 7;9(8):1853. doi: 10.3390/cells9081853.

62. Sulkowski PL et al. 2-Hydroxyglutarate produced by neomorphic IDH mutations suppresses homologous recombination and induces PARP inhibitor sensitivity. Sci Transl Med. 2017 Feb 1;9(375):eaal2463. doi: 10.1126/scitranslmed.aal2463.

63. Sulkowski PL et al. Krebs-cycle-deficient hereditary cancer syndromes are defined by defects in homologous-recombination DNA repair. Nat Genet. 2018 Aug;50(8):1086–1092. doi: 10.1038/s41588-018-0170-4. Epub 2018 Jul 16.

64. Deans AJ et al. DNA interstrand crosslink repair and cancer. Nat Rev Cancer. 2011 Jun 24;11(7):467–80. doi: 10.1038/nrc3088.

65. Ellrott K et al. Scalable Open Science Approach for Mutation Calling of Tumor Exomes Using Multiple Genomic Pipelines. Cell Syst. 2018 Mar 28;6(3):271-281.e7. doi: 10.1016/j.cels.2018.03.002.

66. Takaya H et al. Homologous recombination deficiency status-based classification of high-grade serous ovarian carcinoma. Sci Rep. 2020 Feb 17;10(1):2757. doi: 10.1038/s41598-020-59671-3.

67. Huang KL et al. Pathogenic Germline Variants in 10,389 Adult Cancers. Cell. 2018 Apr 5;173(2):355-370.e14. doi: 10.1016/j.cell.2018.03.039.

68. Wang K et al. PennCNV: an integrated hidden Markov model designed for high-resolution copy number variation detection in whole-genome SNP genotyping data. Genome Res. 2007 Nov;17(11):1665–74. doi: 10.1101/gr.6861907. Epub 2007 Oct 5.

69. Law CW et al. voom: Precision weights unlock linear model analysis tools for RNA-seq read counts. Genome Biol. 2014 Feb 3;15(2):R29. doi: 10.1186/gb-2014-15-2-r29.

70. Wainer Katsir K et al. Human genes escaping X-inactivation revealed by single cell expression data. BMC Genomics. 2019 Mar 12;20(1):201. doi: 10.1186/s12864-019-5507-6.

71. Sztupinszki, Z. et al. Migrating the SNP array-based homologous recombination deficiency measures to next generation sequencing data of breast cancer. npj Breast Cancer 4, 16 (2018).

72. Petljak, M. et al. Characterizing Mutational Signatures in Human Cancer Cell Lines Reveals Episodic APOBEC Mutagenesis. Cell 176, 1282-1294.e20 (2019).

73. Ghandi, M. et al. Next-generation characterization of the Cancer Cell Line Encyclopedia. Nature 569, 503–508 (2019).

