## Supplementary Figures for "The utility of homologous recombination deficiency biomarkers across cancer types"

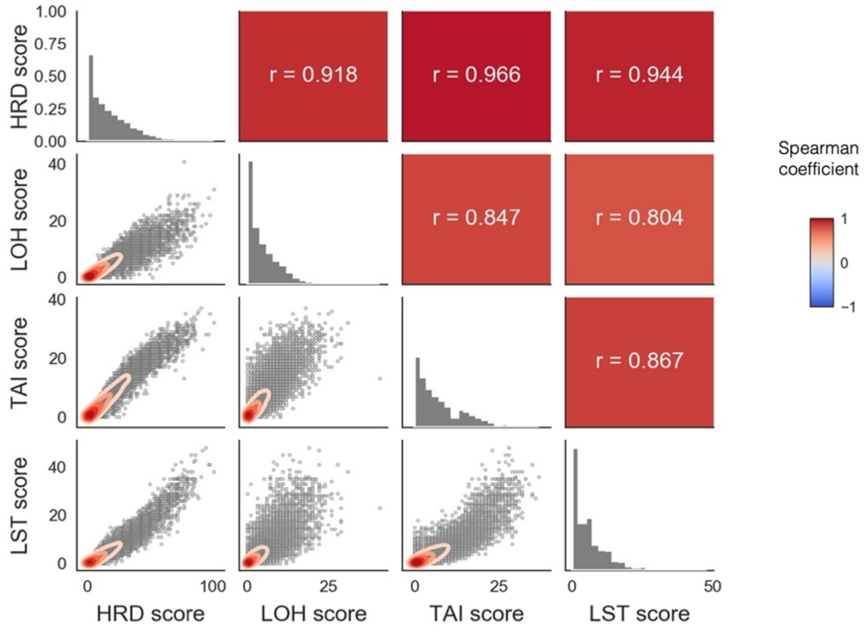


Figure S1. Correlations among LOH, TAI, LST, and HRD scores in all cases.
Lower left panels: scatter and density plots. Upper right panels: Spearman correlation coefficient. Diagonal axis: histograms of each score. All of the Spearman p-values were extremely small ($P < 1\times 10^{-100}$).

A

| Germline | | FACETS | | |
|---|---|---|---|---|
| | | LOH + | LOH - | unknown |
| ASCAT | LOH + | **164** | 0 | **16** |
| | LOH - | 12 | 214 | 8 |
| | unknown | **1** | 1 | 0 |

B

| Somatic | | FACETS | | |
|---|---|---|---|---|
| | | LOH + | LOH - | unknown |
| ASCAT | LOH + | **164** | 5 | **12** |
| | LOH - | 58 | 811 | 65 |
| | unknown | **6** | 8 | 1 |

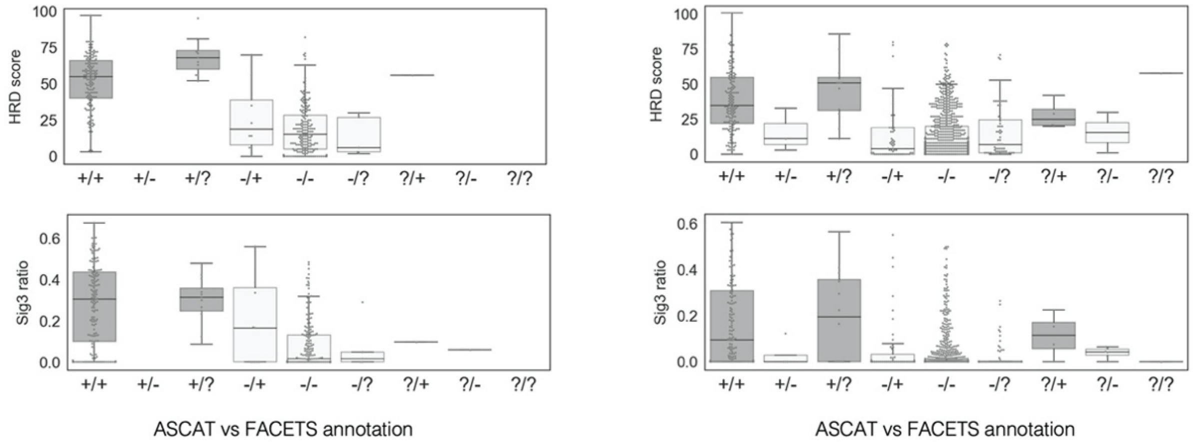


Figure S2. Assessment of the locus-specific LOH.

First, we examined the concordance between the results from ASCAT and FACET for the presence of locus-specific LOH in each somatic (A) and germline (B) variant in the HRR pathway genes (upper). Excluding the samples annotated as unknown, the consistency of the ASCATS and FACETS results was 96.9% and 93.9% for germline and somatic, respectively. Next, we compared HRD scores and Sig3 ratio among the different classification outcome groups (lower). The variants that were determined to have LOH by both algorithms, or to have LOH in one algorithm and to be unknown in the other, showed higher genomic scar scores than the other groups. From these observations, we determined these variants as those accompanying locus-specific LOH. The lower box and whisker plots showed the HRD score and Sig3 ratio of each variant and group. Plus, minus, and question signs indicate LOH, non-LOH, and unknown annotations, respectively. Variants in the bold boxes in the upper tables and those in the dark gray in the lower box plots were determined to have locus-specific LOH.

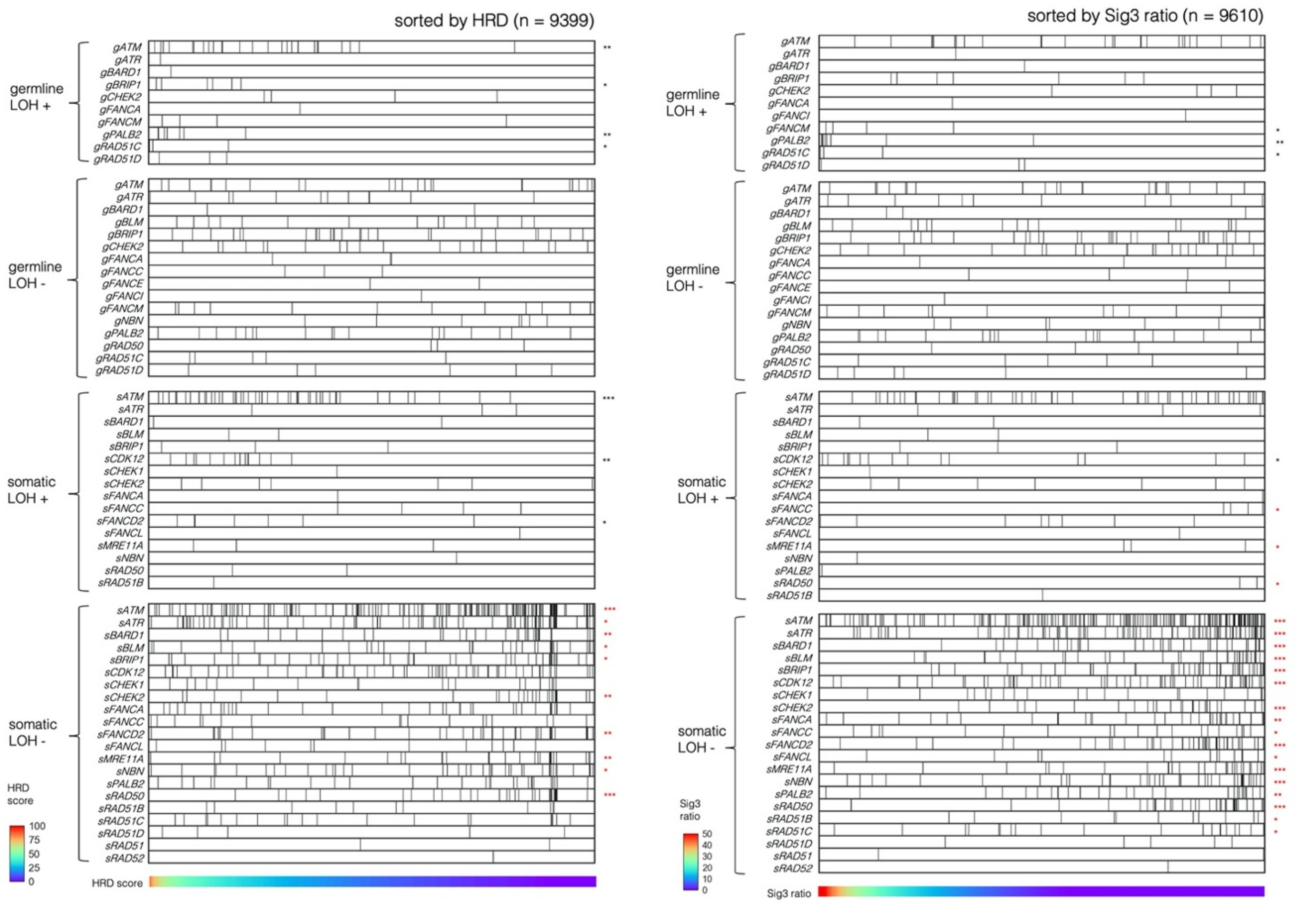


Figure S3. Distribution of germline and somatic mutations with or without locus-specific LOH in HRR pathway genes excluding *BRCA1/2*.

All samples are ordered by HRD score (left) or Sig3 ratio (right), and genes are arranged in alphabetical order. Somatic mutation without LOH was negatively correlated with HRD score and Sig3 ratio. *, **, and *** indicate $P < 0.01$, $P < 1 \times 10^{-4}$, and $P < 1 \times 10^{-6}$ in the Mann–Whitney U test. Asterisks in red stand for a negative correlation with the corresponding value.

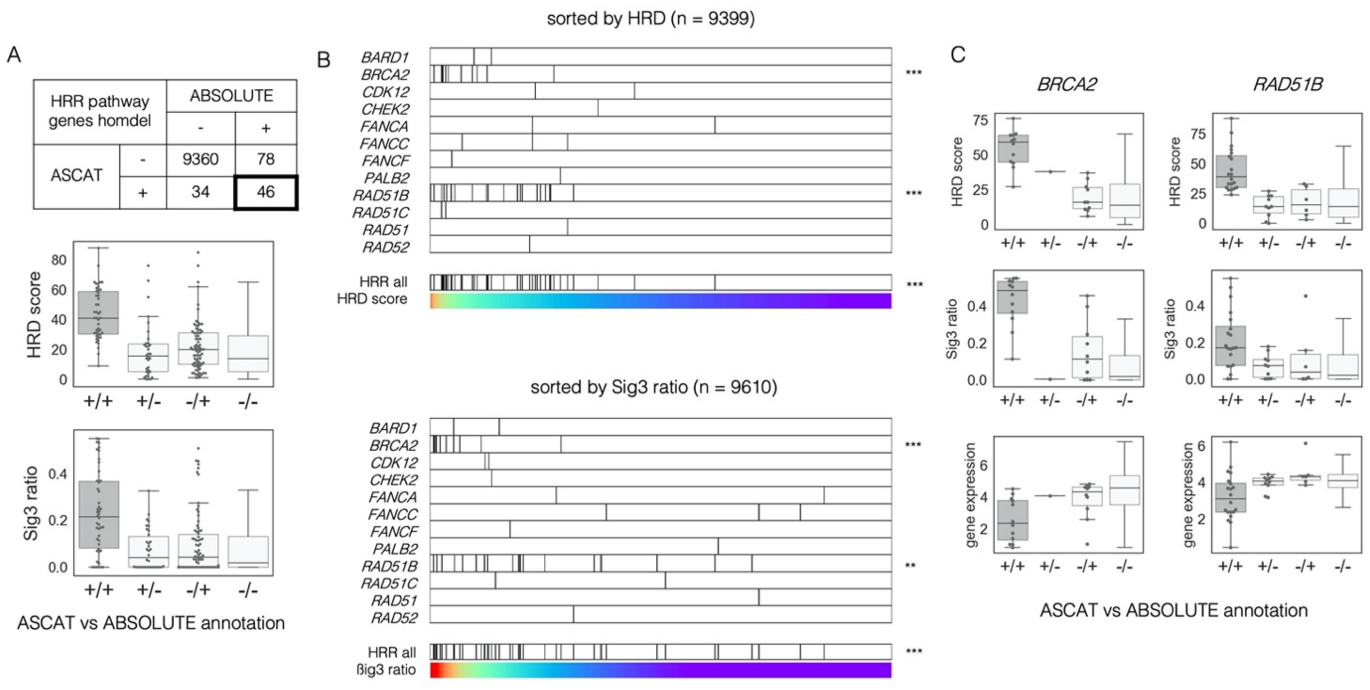


Figure S4. Determination of homozygous deletion.

A) Annotation results for homozygous deletion in the HRR pathway genes from ASCAT and ABSOLUTE were compared. The two results are tabulated in the upper table. HRD score and Sig3 ratio of the cases were compared between each combination of the annotations in the lower box and whisker plots. Plus and minus signs mean having homozygous deletion and not, respectively. Genes in the bold box at the table and dark gray at the box and whisker plots were determined to have homozygous deletion. B) Distribution of homozygous deletion in each HRR pathway gene sorted by HRD score and Sig3 ratio. *, **, and *** indicate $P < 0.01$, $P < 1 \times 10^{-4}$, and $P < 1 \times 10^{-6}$ in the Mann–Whitney U test. C) Comparison of the annotation results of homozygous deletion to HRD score, Sig3 ratio, and mRNA expression in *BRCA2* and *RAD51B*.

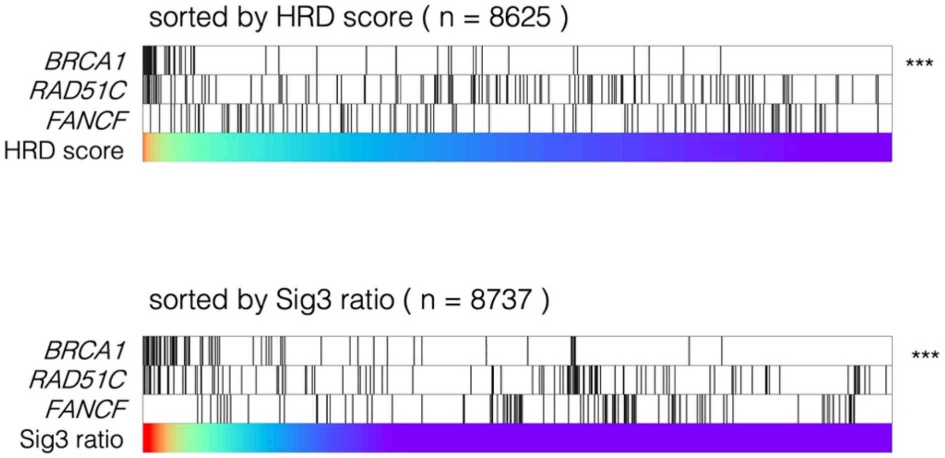


Figure S5. Correlation between promoter methylation-driven gene silencing of HRR pathway genes and genomic scar signatures.

All samples were arranged in order of HRD score (top) and Sig3 ratio (bottom); only *BRCA1* methylation was positively correlated with HRD score and Sig3 ratio. *, **, and *** indicate $P < 0.01$, $P < 1 \times 10^{-4}$, and $P < 1 \times 10^{-6}$ in the Mann–Whitney U test.

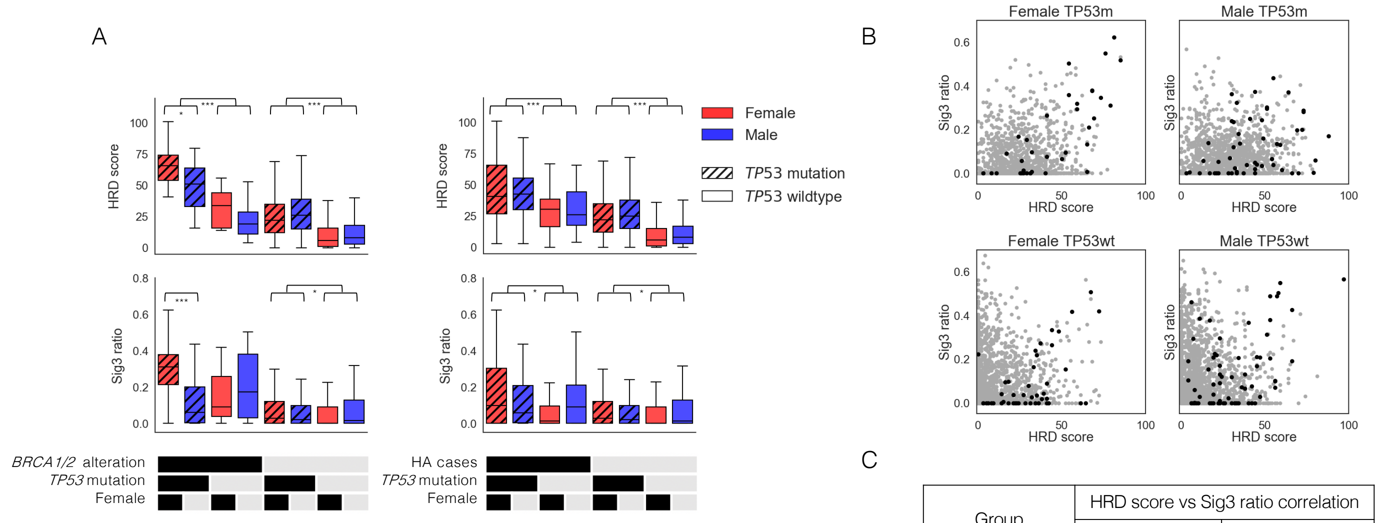


C

| Group | HRD score vs Sig3 ratio correlation | |
|---|---|---|
| | Coefficient | p-value |
| Female TP53m | 0.234 | 6.7e-16 |
| Male TP53m | 0.125 | 2.7e-7 |
| Female TP53wt | 0.062 | 3.1e-3 |
| Male TP53wt | -0.008 | 0.69 |

Figure S6. Differences in genomic scar scores by gender and *TP53* mutation in non-*BRCA*-associated cancers (except ovarian, breast, prostate, and pancreatic cancers).

A) Comparison of HRD score and Sig3 ratio in the eight groups divided by *TP53* mutation, gender, and *BRCA1/2* alterations (including biallelic germline/somatic *BRCA1/2* mutation, plus *BRCA1* methylation, left) or biallelic germline/somatic alteration in all of the HRR pathway genes (right). *, **, and *** indicate $P < 0.05$, $P < 0.01$, and $P < 0.001$ in the Mann–Whitney U test, respectively. B) Comparison of correlation between HRD score and Sig3 ratio and distribution of biallelic HRR pathway gene alterations among the four groups divided by gender and *TP53* mutation. Black dots represent the HRR pathway gene-altered cases and gray represents the other cases. C) Spearman correlation coefficients between HRD score and Sig3 ratio among the four groups. TP53m and TP53wt indicate with and without *TP53* mutation, respectively.

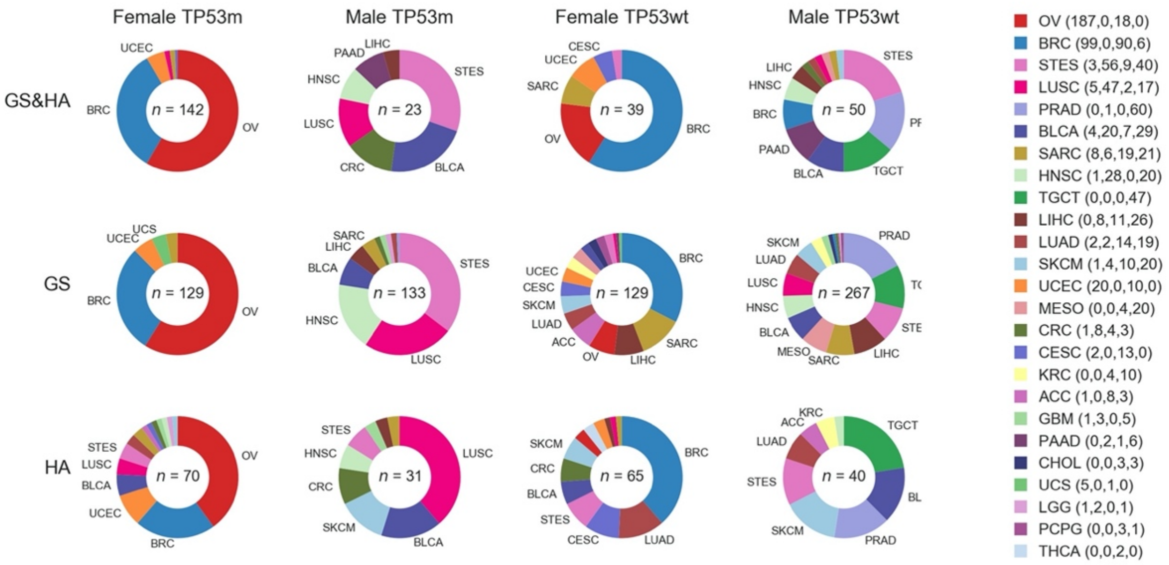


Figure S7. Cancer types and the number of cases assigned to GS cases and/or HA cases in the four groups divided by gender and the presence of *TP53* mutation.

Cancer types that account for more than 3% in each category were labeled.

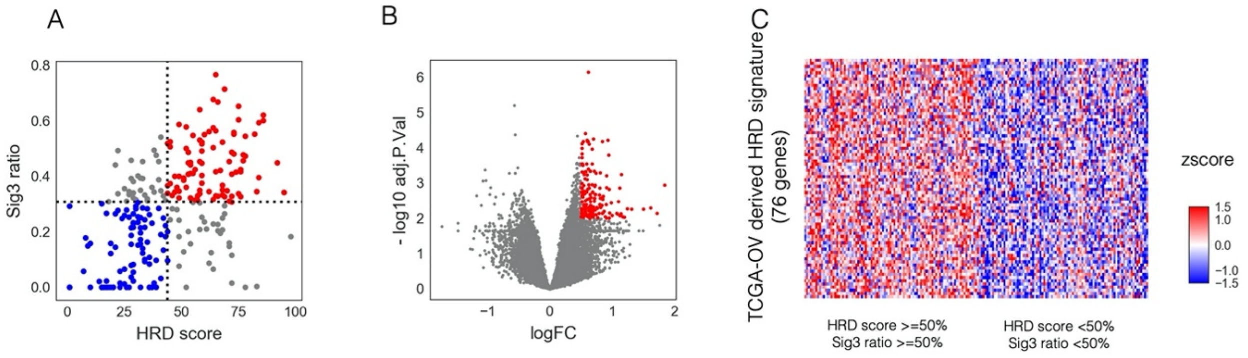


Figure S8. Identification of TCGA OV-derived HRD signature.

(A) Scatter plot of HRD score (X-axis) and Sig3 ratio (Y-axis) for 247 cases of TCGA ovarian cancer. Red dots represent cases with two scores above the 50th percentiles for both, blue dots represent cases below them, and gray dots represent the other cases. B) The result from differentially expressed gene analysis using limma+voom between the above samples by a volcano plot. The dots indicate the genes remaining after filtering with AveExpr > −2. The red dots represent the genes with an adjusted p-value <0.01 and log2 fold change >0.5, namely, TCGA OV-derived HRD signature (see also Supplementary table 4). C) Heatmap of TCGA OV-derived HRD signature gene expression between the above samples. Expression values were log2-transformed and z-scaled for each gene.

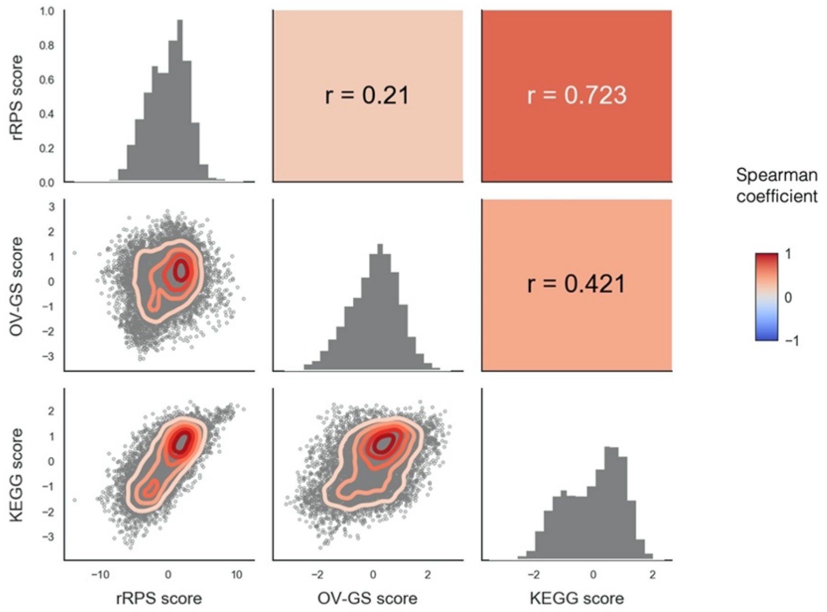


Figure S9. Correlations among rRPS score, OV-GS score, and KEGG score.
Lower left panels: scatter and density plots. Upper right panels: Spearman correlation coefficient. Diagonal axis: histograms of each score. All of the Spearman p-values were extremely small ($P < 1 \times 10^{-92}$).

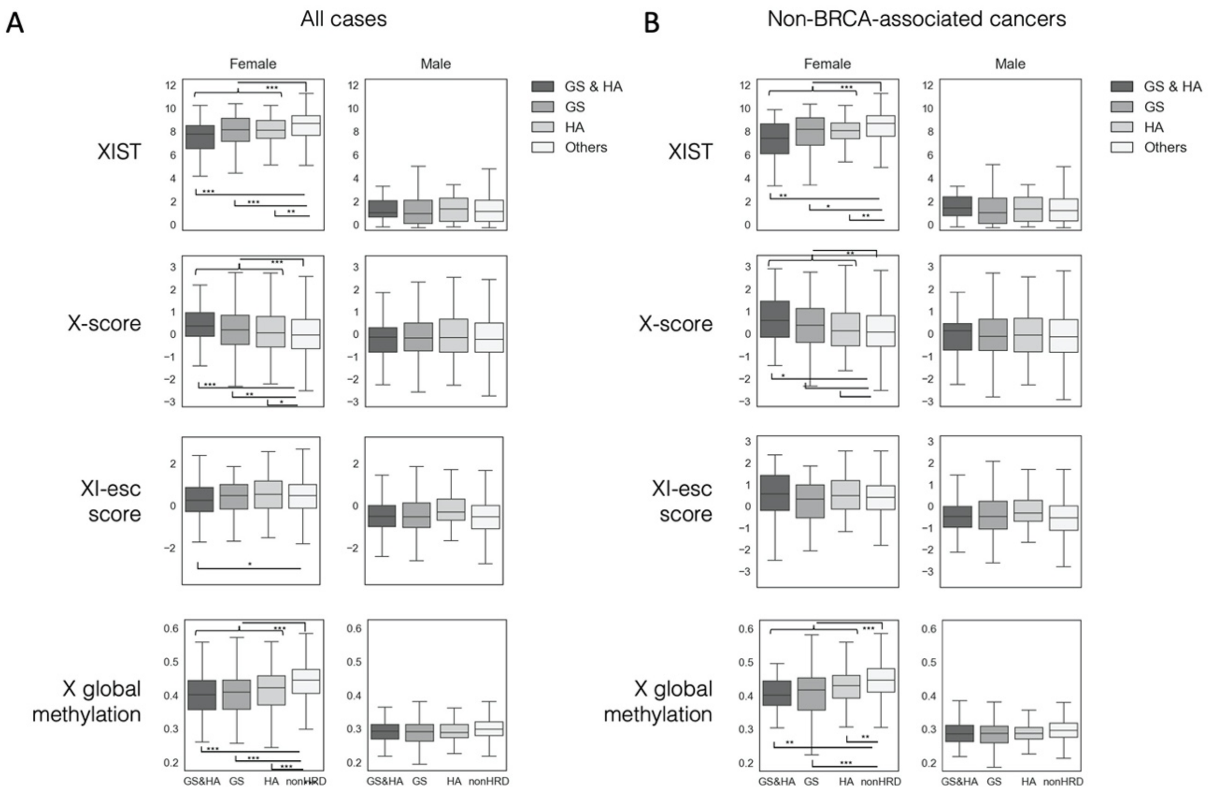


Figure S10. Differences in X chromosome inactivation by gender and HRD status.

Left: in all cases. Right: non-*BRCA*-associated cancers (excluding ovarian, breast, prostate, and pancreatic cancers). The p-values by the Mann–Whitney U test comparing GS and/or HA to non-HRD in all females, all males, non-*BRCA* females, and non-*BRCA* males are $7.2 \times 10^{-18}$, 0.35, $1.2 \times 10^{-5}$, and 0.48 in XIST, $4.7 \times 10^{-8}$, 0.27, $3.8 \times 10^{-3}$, and 0.27 in X score, respectively. The p-values by the Mann–Whitney U test comparing GS and/or HA to non-HRD in all females and non-*BRCA* females are 0.2 and 0.4 in XI-esc score, $1.7 \times 10^{-35}$ and $9.0 \times 10^{-10}$ in X chromosome global methylation, respectively. *, **, and *** indicate $P < 0.05$, $P < 0.01$, and $P < 0.001$ in the Mann–Whitney U test, respectively.

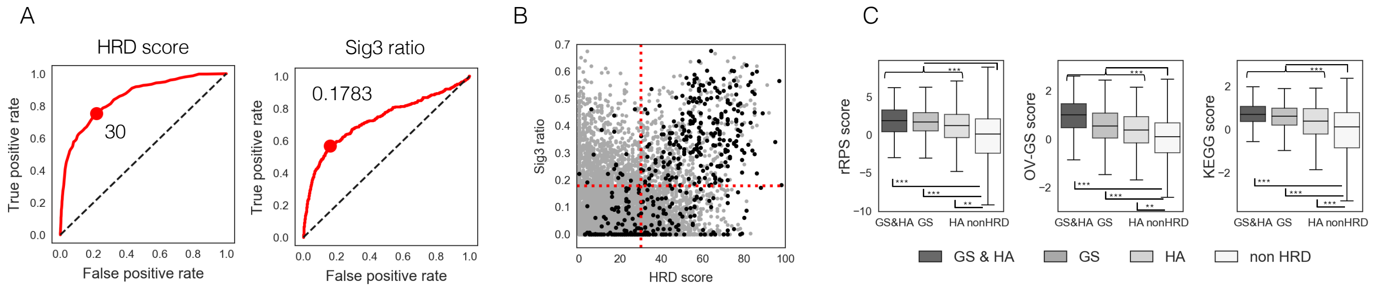


D

| Group | Factor | Number | Univariate Hazard ratio (95%CI) | Univariate p-value | Multivariate Hazard ratio (95%CI) | Multivariate p-value |
|---|---|---|---|---|---|---|
| DNA damaging agent + | Age >= 61 | 1234 (2979) | 1.76 (1.56-1.99) | 1.1e-20 | 1.59 (1.38-1.84) | 3.6e-10 |
| | Male | 1135 (2979) | 1.58 (1.41-1.79) | 3.6e-14 | 1.36 (1.17-1.59) | 6.2e-05 |
| | Stage III/IV | 1240 (2345) | 2.04 (1.76-2.37) | 1.3e-20 | 1.91 (1.64-2.23) | 1.1e-16 |
| | *TP53* mutation | 1585 (2979) | 1.12 (0.99-1.26) | 6.9e-02 | 1.38 (1.18-1.61) | 5.5e-05 |
| | HRD cases | 523 (2979) | 0.80 (0.69-0.94) | 6.0e-03 | 0.83 (0.69-0.99) | 0.043 |
| DNA damaging agent - | Age >= 61 | 3479 (6460) | 2.15 (1.94-2.37) | 2.3e-50 | 1.91 (1.71-2.13) | 5.2e-31 |
| | Male | 3417 (6460) | 1.18 (1.07-1.30) | 5.7e-04 | 1.22 (1.10-1.35) | 1.1e-04 |
| | Stage III/IV | 1743 (5236) | 2.83 (2.56-3.13) | 2.1e-91 | 2.62 (2.37-2.90) | 1.3e-77 |
| | *TP53* mutation | 2089 (6460) | 2.00 (1.82-2.20) | 4.7e-47 | 1.63 (1.47-1.81) | 1.3e-20 |
| | HRD cases | 378 (6460) | 1.57 (1.33-1.87) | 2.2e-07 | 1.37 (1.14-1.64) | 7.7e-04 |

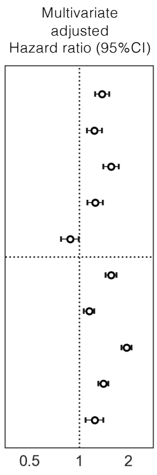


Figure S11. Reanalysis of HRD cases without patient stratification by gender and *TP53* mutation.
A) ROC curves and optimal cutoff values to determine HRR pathway gene alterations by HRD score and Sig3 ratio. B) Correlation between HRD score and Sig3 ratio and distribution of HRR pathway gene alterations in all samples. The red dotted line represents the optimal cutoff values determined in the ROC curves. The black dots represent the HRR pathway gene-altered cases, and the gray dots represented the other cases. C) Comparison of the gene expression-based HRD assessment scores between GS and/or HA and non-HRD cases. The p-values by the Mann–Whitney U test comparing GS and/or HA to non-HRD were $2.6 \times 10^{-52}$ in the rRPS score, $3.6 \times 10^{-46}$ in the OV-GS score, and $4.4 \times 10^{-46}$ in the KEGG score, respectively. *, **, and *** indicate $P < 0.01$, $P < 1 \times 10^{-5}$, and $P < 1 \times 10^{-7}$ in the Mann–Whitney U test. D) Left: univariate and multivariate Cox proportional hazard model analyses with covariates of age, gender, *TP53* mutation, clinical stage, and HRD cases. Right: forest plots showing adjusted hazard ratios with 95%CIs from the multivariate Cox analysis.

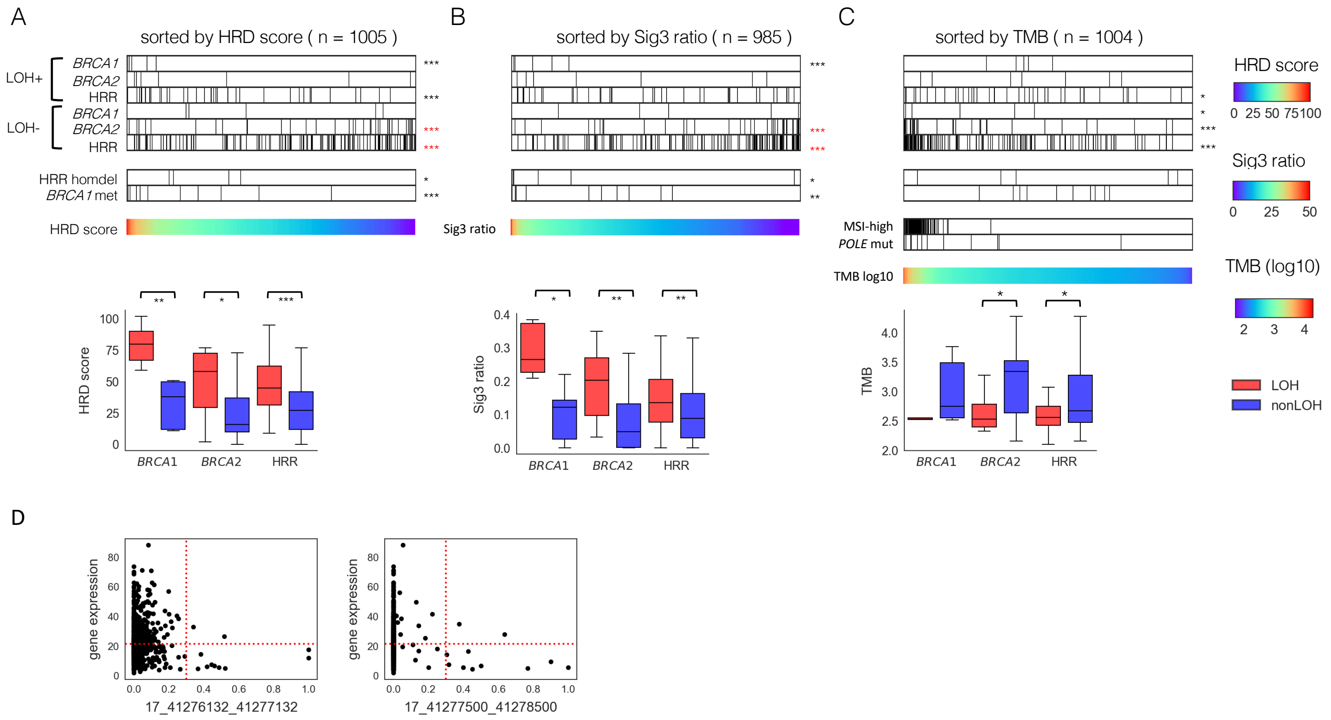


Figure S12. Association between HRR pathway gene alterations and genomic scar scores or tumor mutation burden in CCLE cell lines.

All samples were arranged in order of HRD score (A), Sig3 ratio (B), and TMB (C), respectively. A,B,C) The top panels show the distribution of mutations with and without locus-specific LOH, stratified by *BRCA1*, *BRCA2*, or HRR pathway genes (except for *BRCA1/2*). The second panels show homozygous deletions in HRR pathway genes and *BRCA1* methylation. The five homozygous deletions were *RAD51B, FANCL, RAD51B, BRCA1*, and *FANCC* in order of HRD score. The bottom panels contain the values used for ordering, and samples with MSI-high and somatic POLE mutation in C. The bottom box plots show comparisons of the scores between cases with and without locus-specific LOH in *BRCA1, BRCA2,* or other HRR pathway gene mutations, respectively. *, **, and *** stand for $P < 0.05$, $P < 0.01$, and $P < 0.001$ in the Mann–Whitney U test, respectively. Asterisks in red mean a negative correlation with the corresponding value. D) For BRCA1 methylation, samples with promoter methylation beta values of 0.3 or higher and gene expression below the median of all samples were determined to have methylation-associated silencing.

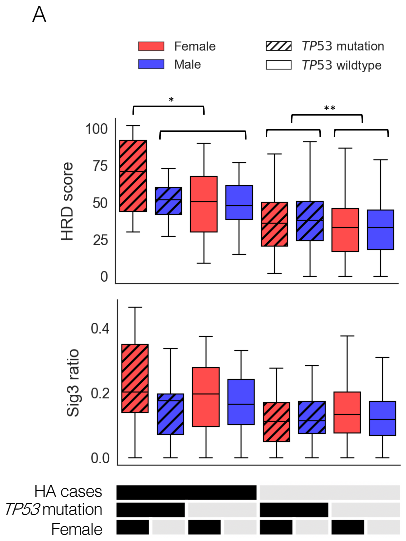


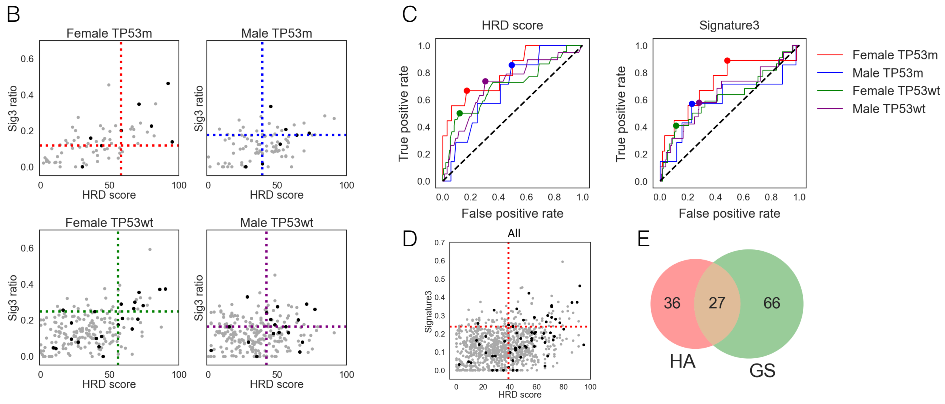


F

| Group | N | HRD score and Sig3 ratio correlation | | HRD score | | Sig3 ratio | |
|---|---|---|---|---|---|---|---|
| | | Coefficient | p-value | Cutoff | AUC (95%CI) | Cutoff | AUC (95%CI) |
| Female TP53m | 73 | 0.613 | 2.2e-8 | 58 | 0.81 (0.63-0.99) | 0.119 | 0.72 (0.53-0.92) |
| Male TP53m | 92 | 0.298 | 4.3e-3 | 39 | 0.69 (0.47-0.82) | 0.176 | 0.60 (0.37-0.83) |
| Female TP53wt | 206 | 0.344 | 4.5e-7 | 56 | 0.70 (0.58-0.83) | 0.249 | 0.62 (0.49-0.76) |
| Male TP53wt | 243 | 0.038 | 0.56 | 42 | 0.71 (0.58-0.85) | 0.165 | 0.64 (0.50-0.78) |

Figure S13. Identification of genomic scar high (GS) cases in CCLE cell lines.

A) Comparison of HRD score and Sig3 ratio in the eight groups divided by *TP53* mutation, gender, and biallelic HRR pathway gene alteration (including BRCA1 methylation). *, **, and *** stand for $P < 0.05$, $P < 0.01$, and $P < 0.001$ in the Mann–Whitney U test, respectively. B) Comparison of correlations between HRD score and Sig3 ratio and distributions of HRR pathway gene-altered (HA) cases among the four groups divided by gender and *TP53* mutation. Black dots represent HA cases and gray dots the other cases. The dotted lines represent the optimal cutoff values determined by the ROC curves in C. C) Differences in ROC curves and optimal cutoff values for HA cases in HRD score and Sig3 ratio among the four groups. D) Correlation between HRD score and Sig3 ratio, the distribution of HA cases, and optimal cutoffs in all samples. E) Venn diagram showing the overlap between the HA and GS cases. F) Spearman correlation coefficients between HRD score and Sig3 ratio, optimal cutoff values, and AUCs on the ROC curves among the four groups. TP53m and TP53wt indicate with and without *TP53* mutation, respectively.
